# Estimating the potential impact of surveillance test-and-treat posts to reduce malaria in border regions in sub-Saharan Africa: a modelling study

**DOI:** 10.1101/2024.06.28.24309631

**Authors:** Hillary M. Topazian, Giovanni D. Charles, Nora Schmit, Matteo Pianella, John M. Marshall, Immo Kleinschmidt, Katharina Hauck, Azra C. Ghani

**Affiliations:** MRC Centre for Global Infectious Disease Analysis, Imperial College London, London, United States; Divisions of Epidemiology and Biostatistics, School of Public Health, University of California, Berkeley, United States; Department of Infectious Disease Epidemiology, London School of Hygiene & Tropical Medicine, London, United Kingdom; Wits Research Institute for Malaria, School of Pathology, Faculty of Health Sciences, University of the Witwatersrand, Johannesburg, Gauteng, South Africa; Southern African Development Community Malaria Elimination Eight Secretariat, Windhoek, Namibia; MRC Centre for Global Infectious Disease Analysis, Jameel Institute, Imperial College London, London, United Kingdom

## Abstract

The last malaria cases in near-elimination settings are often found in international border regions due to the presence of hard-to-reach populations, conflict, uneven intervention coverage, and human migration. Test-and-treat border posts are an under-researched form of active case detection used to interrupt transmission chains between countries. We used an individual-based, mathematical metapopulation model of *P. falciparum* to estimate the effectiveness of border posts on total cases in malaria-endemic sub-Saharan Africa. We estimated that implementation of international border posts across 401 sub-national administrative units would avert a median of 7,173 (IQR: 1,075 to 23,550) cases per unit over a 10-year period and reduce *Pf*PR_2-10_ by a median of 0.21% (IQR: 0.04% to 0.44%). Border posts were most effective in low-transmission settings with high-transmission neighbors. Border posts alone will not allow a country to reach elimination, particularly when considering feasibility and acceptability, but could contribute to broader control packages to targeted populations.

## INTRODUCTION

The World Health Organization (WHO) has selected 25 countries on the fringes of the malaria map to take part in the E-2025 initiative, with the goal of eliminating malaria in these settings by 2025.^1^ However, due to variations in vector habitats, human behavior patterns, intervention coverages, and medical capacities to treat and prevent infection, malaria infection patterns often differ within and between sub-national areas and their nearest neighbors. The result is that countries that have recently eliminated malaria or are on the pathway to elimination often share a border with mid-to high-transmission areas.^1^ Within near-elimination countries, border areas often contain the last malaria cases, due to the presence of remote populations, mobile workers, and/or political complexities limiting the reachability of malaria prevention, diagnosis, and treatment.^2^ Borders also artificially divide transmission foci; malaria intervention coverage may unequal on either side of a border leading to insufficient resources to reduce transmission in a border area.

A key driver of malaria transmission in near-elimination countries is human migration (and to a lesser extent mosquito movement). For example, in South Africa, a large proportion of cases are imported (64.8% in 2019 and 49.1% in 2021) following reductions in indigenous transmission due to malaria control efforts.^3^ Human movement patterns have been characterized using survey data and parasite genetic lineage data have confirmed established transmission chains across international borders, both to adjacent border area villages and across long distances to cities further inland.^4^ Mobile and migrant groups, and even non-travelers who live in communities of individuals with high travel frequencies, may be at an increased risk for infection compared to the general population.^5^ In some instances such as in Mauritius and Armenia, human migration can lead to re-establishment of endemic transmission in an area which has previously eliminated malaria, such as through workers coming in to rebuild after natural disasters, refugees fleeing conflict zones, and military movements.^6^

Two main strategies exist to limit the introduction of new infections from one country to another caused by human movement: A) targeting the “source” population where most infections originate, and B) intervening during migration or shortly after entry to interrupt transmission chains before local onward transmission can occur. Several approaches have been deployed to counteract importation using these two strategies, including forming regional initiatives to fund interventions in source areas,^7^ stationing village malaria workers in hard-to-reach zones to provide better access to diagnosis and treatment, deploying mobile malaria clinics for active case detection, employing test-and-treat to distinct migrant worker populations returning from overseas, and installing screening posts along transport routes to intercept migrants, seasonal workers, and travellers.^8–10^

Setting up border posts is one such strategy for intercepting infections before individuals cross from one country into another. Border posts are a form of active case detection involving parasitological testing and treatment of cases.^11^ Historically, border posts have been added to malaria elimination intervention packages on the China-Myanmar border, the Cambodia-Thailand-Laos borders, Bhutan-India border, and in the Elimination 8 region of southern Africa.^2,12^ Although border post use has been described in these areas, little research exists to quantify the intervention’s effectiveness.^13^ The lack of evidence combined with uncertainty around the feasibility of implementation has led WHO to make a conditional recommendation against routine test-and-treat at points of entry.^11^ More research is needed to determine if border posts can be an effective tool in an elimination setting, and if so, which areas are most suitable for implementation. Mathematical modelling is particularly useful in this instance as measuring the benefits of border posts is nearly impossible to disentangle from the effects of other interventions often packaged alongside.

Here we extend an individual-based, mathematical model of *P. falciparum* to include a metapopulation framework, with the goal of exploring the potential utility of border posts along international borders within sub-Saharan Africa. Our objectives are to 1) estimate the potential impact of border posts in reducing cases of malaria among populations living in border areas, 2) identify characteristics of the sites where the implementation of border posts could be most effective, and 3) determine how their impact depends on coverage indicators. Because countries’ last malaria cases are usually identified in border areas, the WHO recommends addressing border malaria early in the elimination agenda, identifying drivers of transmission and defining appropriate interventions.^2^ Quantifying the effects of border posts and characterizing priority areas for implementation could provide additional evidence for an under-studied malaria control tool, inform the agenda of regional malaria elimination collaborations, and stimulate empirical research.

## RESULTS

We used an individual-based, mathematical metapopulation model of *P. falciparum* to estimate the potential impact of border posts in malaria-endemic sub-Saharan Africa and to identify characteristics of sites most amenable to the intervention. The model was parameterized for 636 first sub-national administrative level units in malaria-endemic sub-Saharan Africa, 401 of which included an international border (**Figure 1A-B**). A total of 44 countries were represented. Parameterization included site-specific epidemiological and historical intervention data from 2000 to 2022 to account for existing regional transmission heterogeneities. Transmission across borders via human movement was simulated for border units in adjacent groups of eight using a gravity model based on destination population size and travel time to capture human movement (**Figure 1C-F**). The gravity model assumes that there is an increased probability of movement towards areas of higher population size and lower travel time. Data were not available to incorporate market centers, road conditions, etc. which may also drive movement in certain areas. Groupings of eight administrative units allowed for capture of malaria trends in border areas, but this analysis did not account for importation of malaria cases from border areas to population centers further inland. The median administrative unit population size (2023) was 989,582 (range: 295 to 48 million) (**Figure S1**) and the median travel time between units was 9 hours (range: 15 minutes to 246 hours) (**Figure S2**). Each unit was represented in a median of 5 unique clusters (range: 1 to 24).

**Figure 1.**
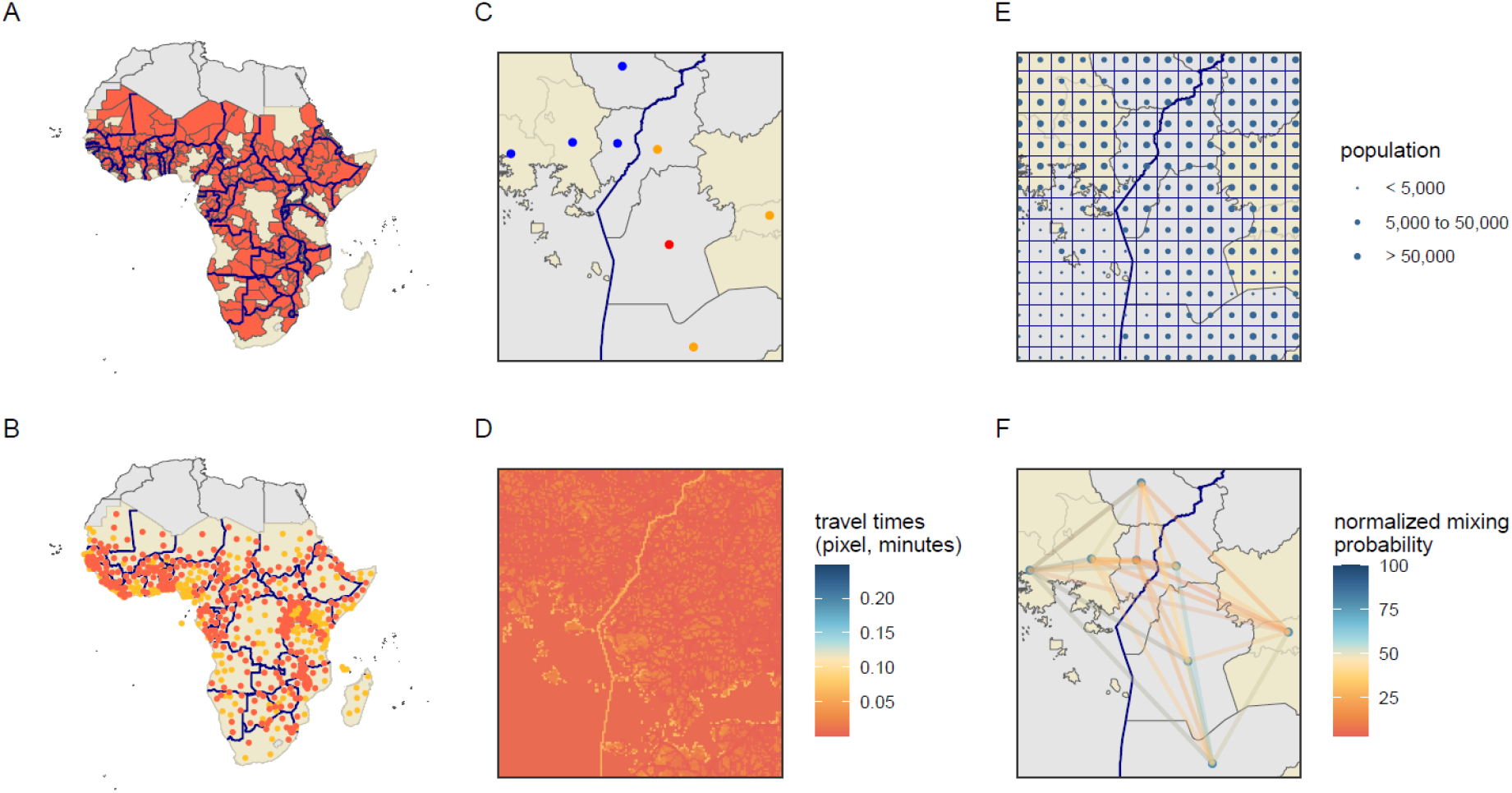
Illustration of approach. Beginning with malaria-endemic countries in sub-Saharan Africa, A) identification of administrative units with an international border (red) and B) identification of administrative unit centroids of border areas (red) and non-border areas (orange). Panels C-F show an example cluster with the international border indicated with a bolded line. Gray polygons indicate border administrative units, yellow polygons indicate non-border units. C) Selection of seed point (red), three nearest neighbors within the same country (orange), and four nearest neighbors outside of the country (blue). D) Travel time raster developed by the Malaria Atlas Project,^14^ each pixel represents the travel time to cross the pixel in minutes with an additional time penalty added at the border. E) Population sizes taken from the cell centroids of a 0.1×0.1 degree grid surface used in partnership with travel times and trip durations to calculate the F) normalized mixing probabilities between units using a gravity model, aggregated from the grid to the administrative unit level.

Simulations of border posts assuming that 80% of people who cross borders undergo rapid diagnostic testing (RDT) and that treatment of positive cases with artemether-lumefantrine is 95% effective, averted a median of 7,173 (IQR: 1,075 to 23,550) cases per administrative unit in all border seed points over a 10-year period (**Figure 2A**). When looking at relative impact, border posts resulted in a median 0.21% decrease (IQR: 0.04% to 0.44%) in *P. falciparum* prevalence among 2–10-year-olds (*Pf*PR_2-10_) from baseline to post-intervention period (**Figure 2B**). Accounting for the variable sensitivity of RDTs, particularly low sensitivity at low *Pf*PR_2-10_ values, a median of 14 (IQR: 5 to 82) people would need to be screened at a border post to prevent one case, across all border seed points (**Figure 3A**). Border posts appeared to be most effective in low transmission areas with high transmission neighbors, similar to areas along the Kenya-Ethiopia-South Sudan borders, Rwanda, and western Côte d’Ivoire regions of sub-Saharan Africa. Due to a lack of sub-national data on human movement patterns, and an optimistic, universally applied border post coverage value of 80%, results should not be interpreted as recommendations for specific countries or sub-national areas.

**Figure 2.**
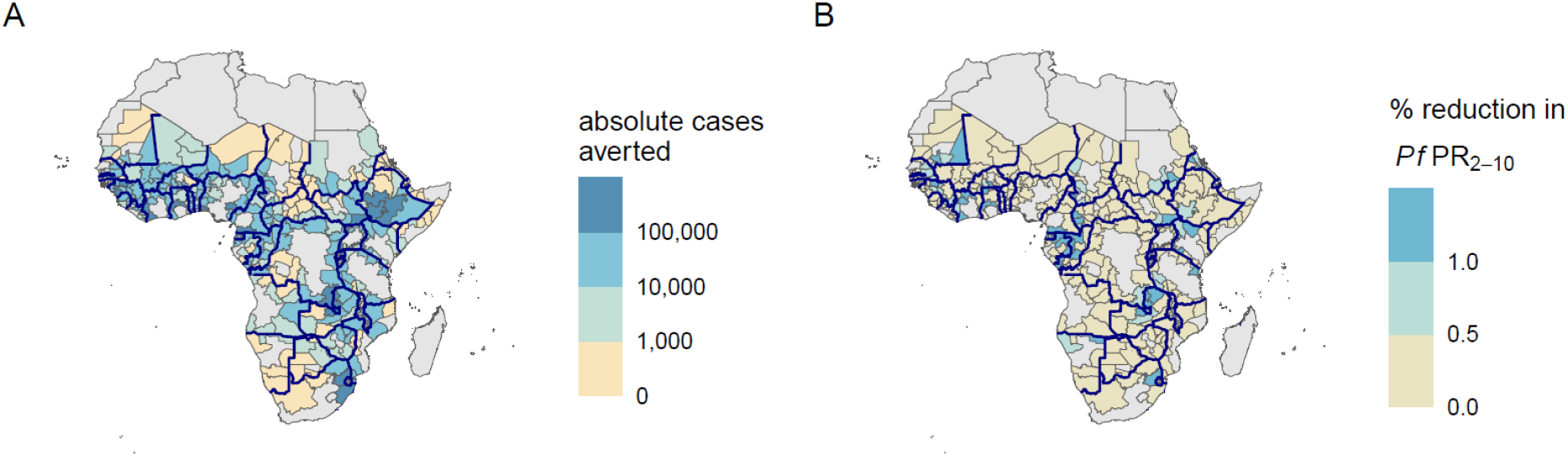
Effects of border post simulations with 80% coverage of rapid diagnostic testing over the 10-year intervention period relative to no intervention. Administrative units are parameterized with unit-specific epidemiological and historical intervention data. Colors represent median values out of 50 unique model parameter draws. Panel A represents the absolute number of cases averted in each unit when it was the seed point for the cluster. In panel B, each unit represents the percent reduction in *Pf*PR_2-10_ from before to after the border post intervention. Bold lines indicate boundaries between countries included in the analysis. All units with outcomes slightly less than 0 due to model stochasticity at extremely low values of *Pf*PR_2-10_ were set at 0.

**Figure 3.**
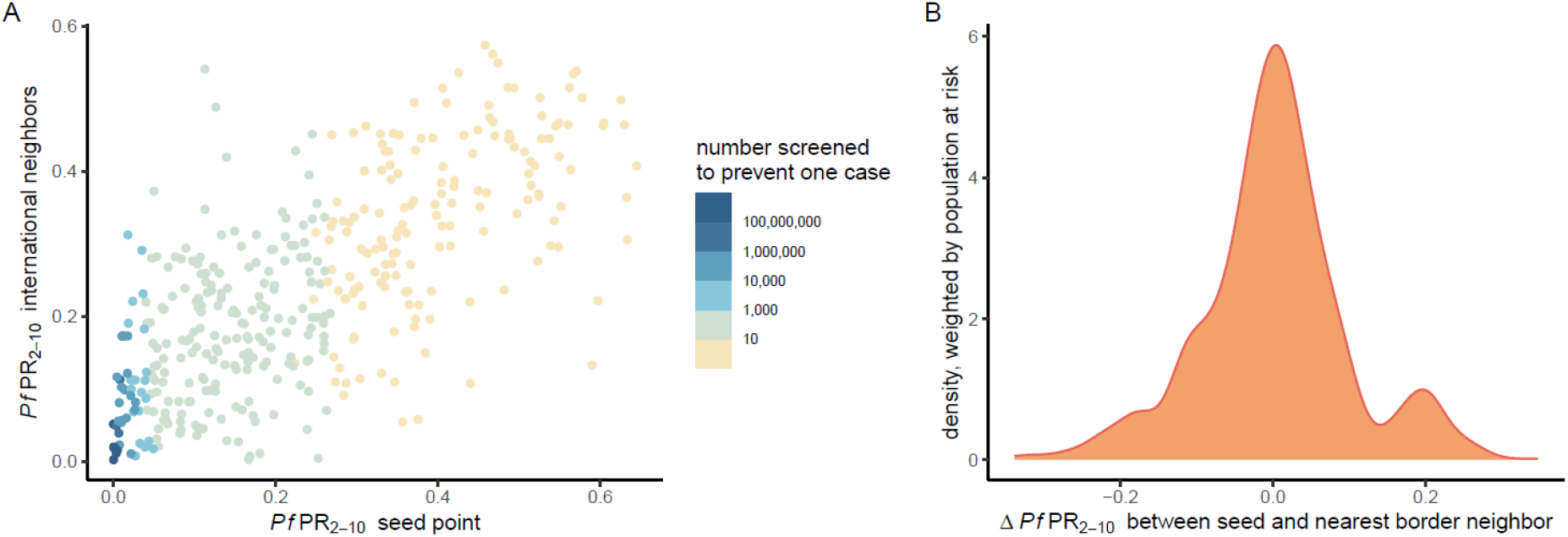
Outcomes by prevalence differences between neighbors. A) The number of people who need to be screened to prevent one case per cluster via rapid diagnostic test. Each point represents the median cluster value of 50 parameter draws, classified into the *Pf*PR_2-10_ of the seed point vs. the *Pf*PR_2-10_ of the four international neighbors in the cluster. B) Density plot of the population at risk of *P. falciparum* by the difference in *Pf*PR_2-10_ between each seed point and its nearest border neighbor.

To further investigate the drivers behind border post effectiveness, two generic case studies were performed to assess the effects of *Pf*PR_2-10_ and intervention coverage on cases averted. The first case study, examining the effectiveness of border posts by *Pf*PR_2-10_ using a 2-unit model, showed that border posts reduced the highest percentage of cases in scenarios where a high transmission unit bordered a low-transmission unit (**Figure 4A**). *Pf*PR_2-10_ units in low-transmission settings of 5% to 10%, paired with high-transmission *Pf*PR_2-10_ units of 50% to 80% resulted in 4.0% to 13.9% of cases averted in low-transmission units in the total population over 10 years. These scenarios are similar to the above-mentioned regions in sub-Saharan Africa, although border neighbors tend to have smaller transmission differences (**Figure 3B**). Border posts had essentially no effect when *Pf*PR_2-10_ combinations of low-transmission units were paired with other low-transmission units, due to the low number of infections mixing and the low sensitivity of RDTs. When border posts were added to the model with testing through polymerase chain reaction (PCR) instead of RDT (representing 100% sensitivity and specificity to infections), results showed similar trends, with a higher proportion of cases averted ranging from 9.5% to 22.9% when low-transmission units of *Pf*PR_2-10_ 5% to 10% were paired with high-transmission units of 50% to 80% (**Figure S3**). A higher proportion of cases were detected by RDT when examining movement from high-transmission areas vs. low-transmission areas, ranging from a median of 25.5% (IQR: 24.8% to 27.4%) across 2-units of *Pf*PR_2-10_ 5% to a median of 96.2% (IQR: 95.3% to 97.4%) across two-units of *Pf*PR_2-10_ 80% (**Figure 4B**).

**Figure 4.**
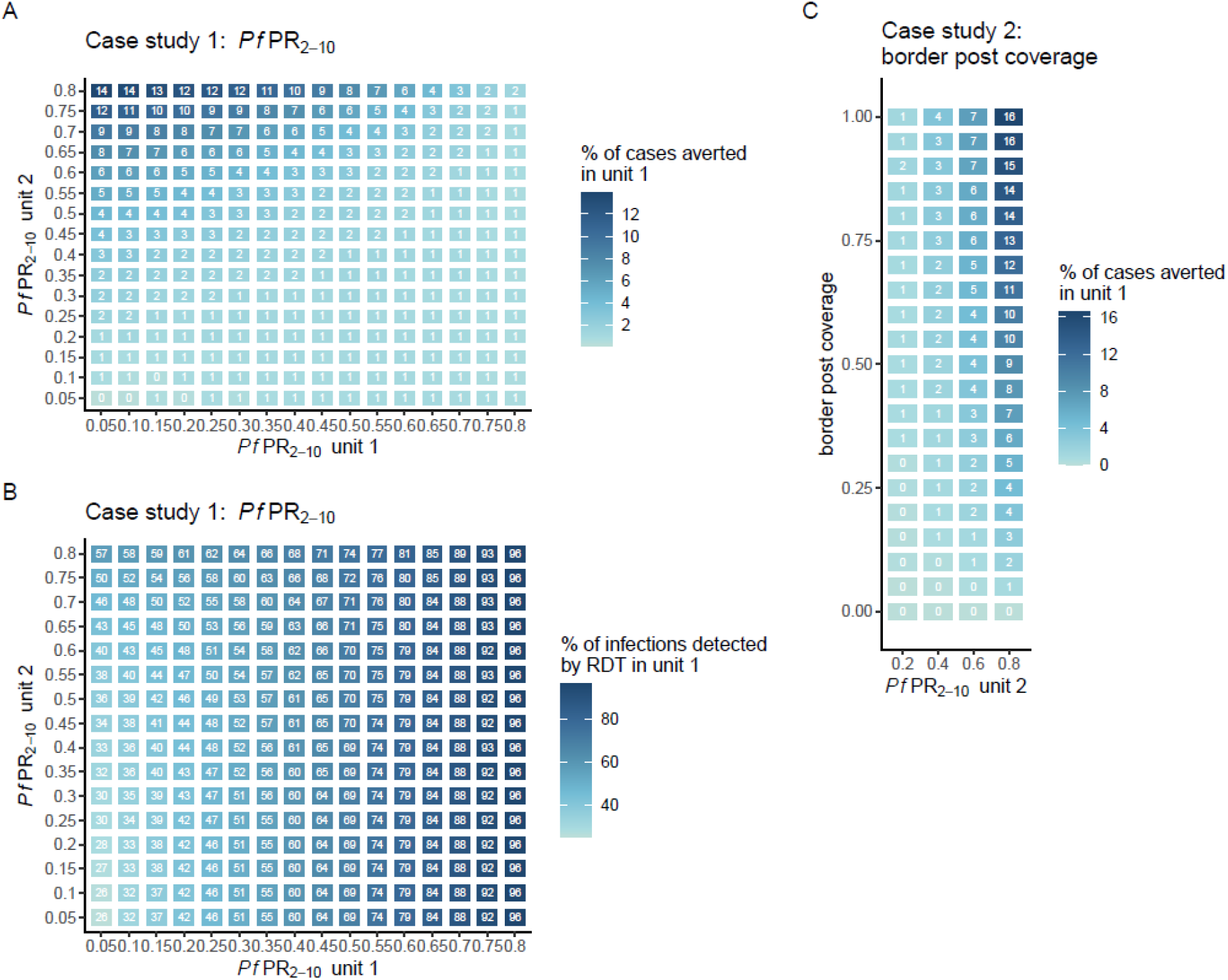
Measuring the sensitivity of outcomes to *Pf*PR_2-10_ and border post coverage. Case study 1 examines the effects of A) *Pf*PR_2-10_ on the percent of cases averted and B) *Pf*PR_2-10_ on the percent of infections detected by RDT. Case study 2 examines the effects of C) border post coverage on the percent of cases averted in the total population using a 2-unit metapopulation model over a 10-year period. Colors represent median values out of 50 unique parameter draws. The model assumes that 95% of mixing occurs within a unit and 5% of mixing occurs between units. Note: RDT = rapid diagnostic test.

Our second case study, examining the effectiveness of border posts by the percentage of transmission captured by the test-and-treat structure using a 2-unit model, demonstrated that higher border post coverage led to a higher percentage of cases averted, but only when a low transmission setting (*Pf*PR_2-10_ 10%) was paired with a medium to high *Pf*PR_2-10_ (60% or 80%) (**Figure 4C**). Values ranged from <1% cases averted when border post coverage was <70% when a *Pf*PR_2-10_ 10% unit was bordering a *Pf*PR_2-10_ 20% unit to a maximum of 16.5% when a *Pf*PR_2-10_ 10% unit was bordering a *Pf*PR_2-10_ 80% unit with 100% border post coverage.

## DISCUSSION

Modelling the effectiveness of border post interventions across 401 uniquely parameterized international border areas in sub-Saharan Africa resulted in a range of outcomes: a median of 7,173 (IQR: 1,075 to 23,550) cases averted per unit and a median 0.21% decrease (IQR: 0.04% to 0.44%) in *Pf*PR_2-10_. Case studies resulted in two general findings. First, that the difference in *Pf*PR_2-10_ values on either side of a border has a large effect on the potential number of cases averted by the intervention. The highest relative cases averted occurred in near-elimination settings which border a high transmission neighbor, such as *Pf*PR_2-10_ units of 50% to 80% paired with *Pf*PR_2-10_ units of 5% to 10%, which averted 4.0% to 13.9% of cases due to the high probability of intercepted travelers carrying infection. Border posts are unlikely to be an effective solution in near-elimination settings which border other low-transmission areas due to the infrequent likelihood of cases and low sensitivity of RDTs since many infected travelers may have undetectable infections due to low-parasitemia, and screening the large number of people that need to be tested to detect a single infection is difficult to achieve. Second, that the proportion of all travelers screened at a border post has influence on overall cases averted in settings where near-elimination units border high-transmission units. When implementing border posts in appropriate settings, being able to cover highly frequented travel routes or specific populations at high risk of transmitting infection will be important to effectively halt cross-border transmission. Although border posts are unlikely to allow a country to reach elimination in isolation, they can contribute to elimination efforts in border areas as part of a wider package of regional surveillance, control, and health system strengthening.

Border posts have been used historically in elimination settings in Asia on the China-Myanmar border and the Cambodia-Thailand-Laos borders as part of broader intervention packages.^2^ Only one program implementing border posts has been described in sub-Saharan Africa, which took place in the Elimination 8 region of southern Africa, involving 46 malaria health posts stationed along the five international borders in the region from 2016-2018.^15^ These locations appeared fit for border post implementation at the time, given the presence of a near-elimination country paired with higher transmission neighbors, as seen in our case studies. In the Elimination 8 region it was estimated that mobile and static malaria health units contributed to an estimated 30% reduction in malaria incidence in border areas among other activities, although this decrease was not enough to eliminate malaria.^15^ Mathematical modelling in the region corroborates these findings, showing that focalized screen-and-treat border interventions could have large but short-lived effects, would need to be continuously implemented to prevent renewed transmission, and would be insufficient to eliminate local infection at less than 100% coverage.^16^ Screen and treat methods could also be effective in island situations where there is often a large difference in transmission between the elimination island and the nearest mainland location from which most travelers originate.^5,17^

These findings highlight the importance of regional cooperation to supplement border post activities through initiatives such as regional resource sharing,^7^ working with private organizations to screen at-risk occupational groups,^18^ improving health systems in low-resourced areas,^7^ and coordinating vector control campaigns across borders.^19^ Implementing a package of interventions is particularly important as it is unknown what coverage can be feasibly achieved with border posts given the number of informal border crossings in most locations. In places where malaria control decision-makers face national-level political complexities, local collaboration between bordering administrative units could be a more important near-term goal than national-level collaboration.^2^ Border posts, while potentially reducing the rate of importation into countries, cannot take the place of ensuring quality implementation and high coverage of malaria interventions by national and subnational malaria programs.

National malaria control programs have formed regional consortiums to tackle malaria control across the globe, spanning from the Elimination of Malaria group in Mesoamerica and Hispaniola, to the Malaria-Free Arabian Peninsula Initiative, African Leaders Malaria Alliance, and the Asia Pacific Malaria Elimination Network, among others.^20^ International funders are supportive of these regional partnerships, with the WHO Global Technical Strategy for Malaria 2016-2030 including a goal to “deepen regional collaboration.”^21^ However, the success of regional initiatives requires external funding. The Lubombo Spatial Development Initiative, for example, was incredibly successful in reducing malaria along border areas of South Africa, Swaziland, and Mozambique,^22^ but after the closure of the program due to a lack of financial resources, malaria rebounded across all three countries.^23^ The Global Fund does invest in a few key multicounty priorities, including 20 million allocated for malaria elimination in Southern Africa and 120 million for drug resistance in the Greater Mekong Sub-region,^24^ but generally the international aid structure currently contains little to accommodate regional proposals in addition to country-specific projects.^20^

The feasibility of establishing border posts and acceptance by the target population is also a significant concern. Set-up of cross-border collaboration test-and-treat methods to targeted groups has been feasible in French Guiana, Suriname, and Brazil through distribution of self-test and treatment kits to mobile gold miner populations.^25^ However, only a small number of studies have published data on static border post user-acceptance to wider traveler populations with mixed results. In Cambodia, 22% of approached travelers refused to participate in the border post intervention due to a lack of time, a perception of no malaria risk, fear of blood draw, and language or cultural barriers.^26^ Alternatively, focus groups regarding a border post in the Solomon Islands indicated high acceptance of test-and-treat, suggesting that in some settings mandatory testing before travel by ship may be feasible if backed up by legislation to empower health workers and reduce noncompliance.^27^ Like coverage, the sensitivity of the diagnostic used was found to be a driver of border post effectiveness and the number of cases detected in our study; PCR has a much lower limit of detection compared to RDTs, but processing time can take hours rather than minutes and the technology may not be feasible to implement within a point-of-care design.^28–30^ If it is not feasible to screen a large proportion of individuals crossing a border, targeting high-risk mobile populations may be a better approach. Plantations in Malaysia have worked with the Malaria Control Programme to screen new workers for malaria upon arrival, many of whom are foreign migrants,^18^ and other countries in Asia have set up programs to screen returning UN peacekeepers and military members.^31^

The primary limitation to this analysis is the inability to capture border post intervention costs, including those related to infrastructure, human resources, and diagnostic and treatment supply chains, all of which are likely to vary between individual settings. We have presented a comparison of the number of people needed to test to prevent one case to represent a measure of resource effectiveness, but the societal value of setting-up and running border posts, like any intervention, will depend on the cost-effectiveness and affordability of implementation relative to other tools such as targeted vector control or regional resource sharing to reduce incidence in “source” populations. Border posts target a much smaller but higher-risk population than general mass drug administration or vector control of large geographic areas, potentially leading to lower costs, but the expense of set-up and maintenance could vary widely depending on existing infrastructure. Additional sub-national modelling will be necessary to inform country decision making. Sensitivity to the resolution of analysis could also be further explored. In this study, although the gravity model was created using a 0.1×0.1 degree grid, historical intervention use and *Pf*PR_2-10_ was parameterized at the first administrative unit level which is quite broad; country-specific work using smaller geographical units (such as level 2 units) representing heterogeneities in *Pf*PR_2-10_ and intervention use could better inform border malaria outcomes. Our analysis is also limited by a lack of country-specific data on human movement, requiring the use of a gravity model and travel time friction surface to build mixing matrices, with between-country movement and within-country movement modelled the same way. There are no data available to inform the percentage of travelers across international borders which would be able to be captured by a border post intervention, and this is likely to be country-specific and depend on the route of travel (walking paths through forests, paved roads, boat, air). In sub-Saharan Africa borders are often porous, making it likely that the effectiveness of border interventions will in practice be much lower than the 80% coverage assumed in this study. Information on where travelers cross the border and the estimated proportion of travelers able to be tested and treated by targeting key routes will be important to generate more accurate, country-specific model runs.

Despite these limitations and a lack of data on costs and feasibility, border posts have the potential to contribute to malaria control beyond the treatment of infected individuals. In the Greater Mekong Subregion, border posts have been used to monitor changes in artemisinin resistant parasites flowing across countries, and to characterize the level of malaria importation stemming from asymptomatic vs. symptomatic individuals.^26^ It is possible that border posts can fill gaps where passive case detection of mobile populations through routine health systems is ineffective; in north-eastern Cambodia, mobile malaria workers near the border contributed to 45% of all testing and detected 39% of all cases registered in border areas.^32^ Bhutan has integrated malaria screening alongside HIV, tuberculosis, and COVID-19 at border towns for foreign workers entering the country and Timor-Leste has integrated malaria interventions into those already existing for dengue, making border interventions more cost effective by targeting a broader range of infectious diseases, and more sustainable as fewer infections are picked up as malaria transmission declines.^12^

Considering the ambitious identification of 25 countries with the potential to eliminate malaria by 2025 and the goal of eliminating malaria in at least 20 countries by 2025,^1^ the global community must encourage regional cooperation and the evaluation of strategies targeted towards border malaria. Border posts could be one effective option to address cross-border transmission in near-elimination areas with higher transmission neighbors. Although the effectiveness of border posts will ultimately depend on the percentage of travelers captured by the intervention, feasibility, and cost-effectiveness, they can also contribute to wider health benefits for the target population when coupled with other aspects of routine care or screening for additional infectious diseases. Future modelling work should assess the implementation of border posts compared to other forms of regional cooperation such as resource sharing and synchronizing vector control campaigns and investigate the role of border posts in settings where a large proportion of malaria cases in the eliminating country are imported cases from neighboring districts.

## METHODS

### Individual-based model

Modelling was performed using *malariasimulation* (*v1*.*6*.*0*; Charles G, et al. 2023), an open-source individual-based mathematical model of *P. falciparum* in R 4.3.2 (R Core Team, 2023). The model has been previously parameterized by fitting to age-stratified severe disease, clinical disease, and parasite prevalence data across sub-Saharan Africa.^33,34^ *malariasimulation* incorporates variations in vector species-specific biting rates, population age-structures, adaptive immunity, seasonality, and intervention usage. In the model, individuals enter at birth and become susceptible to *P. falciparum* infection over time as maternally acquired immunity fades. Individuals become infected with *P. falciparum* with an age-based probability, developing either asymptomatic infection or clinical disease, with a proportion of clinically diseased individuals developing severe disease. Mosquito vectors are modelled compartmentally, and mosquitoes become infectious through biting an infected human. Individual human immunity includes maternal antibodies at birth, pre-erythrocytic (anti-infection) immunity, blood stage (anti-parasitic) immunity, and infection detection immunity which are functions of age and previous exposure to infection. Individual level biting rates are assumed to be heterogeneous in the population.

Anti-malarial interventions incorporated in the model include treatment, insecticide-treated nets (ITNs), indoor residual spraying (IRS), seasonal malaria chemoprevention (SMC), and malaria vaccines, allowing for modelling of historical intervention coverages in specific settings. Treatment clears infection from individuals experiencing clinical disease and provides a drug-dependent partial protection from repeat infection which wanes following a Weibull survival curve. ITNs are implemented by reducing female mosquito attempts to feed and by increasing the probabilities of these mosquitoes being repelled or killed. ITN efficacy is dependent on the type of insecticide used, the level of insecticide-resistance specified, and the age of the net.^35^ Administration of SMC clears existing infection with a drug-dependent probability and provides a period of temporary prophylaxis against re-infection. Malaria vaccine efficacy reduces the probability of infection following administration of the primary doses and follows a biphasic model with short and long lived anti-circumsporozoite protein antibody decay dynamics. RTS,S vaccine parameters were previously fit to data from a multi-site Phase III randomized controlled trial.^36^

Additional details can be found in the Supplementary Information under Technical Methods. Functions and documentation for *malariasimulation* are open source and can be found at: https://github.com/mrc-ide/malariasimulation.

### Metapopulation model

The metapopulation component of *malariasimulation* allows for multiple, simultaneous, interconnected model runs, with each run or “unit” uniquely parameterized for a given setting. Rather than model the movement of individual humans between units, *malariasimulation* simulates movement and spatial interconnectedness by allowing the malaria transmission levels of one unit to influence the malaria transmission levels of neighboring units with a user-specified probability. Malaria transmission levels are captured via the entomological inoculation rate (EIR), the number of infectious mosquito bites per person per day, and the force of infection on mosquitoes (FOIM), the rate of infection acquired by mosquitoes from infectious humans. The probability of influence that one unit exerts on a neighboring unit is drawn from a user-specified mixing matrix, where each row indicates the primary unit, and each column indicates the secondary connected units that may influence transmission within the primary unit. Each element of the matrix can vary between 0 (no mixing) and 1 (representing fully random mixing between two units). An illustrative example of mixing patterns and the resulting effects on malaria outcomes is shown in **Figure S4**.

The probability of human movement between primary and secondary units was calculated using a previously established gravity movement model fit to data from travel surveys in Burkina Faso, Mali, Tanzania, and Zambia.^37^ The model estimates the probability of travel based on destination population size and the travel time between origin and destination population-weighted centroids, with larger destination population and shorter travel times corresponding to higher probabilities. To translate these into connectivity between primary and secondary units we used gravity model estimates to calculate the bi-directional interactions between all cells of a 0.1×0.1 degree grid overlaid on top of the administrative units with these then aggregated by administrative unit. The trip duration estimates were fit using commune- and ward-level data from the same surveys mentioned previously.^38^ The overall mixing matrices were calculated by combining information on (a) the estimated bi-directional travel between administrative units; (b) the trip duration; and (c) the probability of travel, estimated from Demographic and Health Survey data on the number of trips away from home for one or more nights in the last year.^39^ We assumed that between-country and within-country movement patterns were the same (i.e. that borders did not affect movement).

Additional details can be found in the Supplementary Information under Technical Methods.

### Site selection and parameterization

The primary analysis consists of 33-year model runs (representing years 2000 – 2032) for 636 sites, representing all first sub-national administrative level units in malaria-endemic sub-Saharan Africa. 401 of these units include an international border and were the main focus of the analysis (**Figures 1A, 1B**). Islands were excluded as these units did not include an international land border.

Each unit was parameterized using GADM (*v4*.*0*) administrative boundaries,^40^ and site-specific files from the *foresite* (v.0.1.0; Winskill P, 2023) and *site* (v.0.2.2; Winskill P, 2023) packages. Units were characterized from years 2000 to 2022 using data from WorldPop population counts,^41^ World Malaria Report cases and deaths,^21^ Malaria Atlas Project *Pf*PR_2-10_ estimates,^42^ Malaria Atlas Project vector species abundance and distribution,^43^ and Malaria Atlas Project^44^ and the DHS StatCompiler^45^ estimates of historical intervention coverage such as ITN use, IRS, SMC, RTS,S, and treatment. SMC coverage estimates were taken from ACCESS-SMC^46^ and SMC Alliance,^47^ and RTS,S coverage was drawn from the Malaria Vaccine Implementation Programme.^48^ Treatment was categorized as artemether-lumefantrine (an artemisinin combination therapy currently recommended as a first-line treatment)^3^ or sulphadoxine-pyrimethamine (used historically). ITNs were classified as pyrethroid, pyrethroid + PBO, or pytrethroid + pyrrole with setting-specific estimated pyrethroid insecticide resistance levels. IRS was classified using a variety of insecticide options, with the assumption that a DDT-type insecticide was used prior to 2017 and an actellic-like insecticide was used post 2017. All SMC interventions are assigned the drug sulfadoxine-pyrimethamine + amodiaquine. A standard demography profile corresponding to the population age structure in sub-Saharan Africa in 2021 was used across all model runs.^49^ Seasonality profiles for each unit, which remained static across years, were created using *umbrella* (*v0*.*1*.*4*; Winskill P, 2021) which constructs a Fourier series model using CHIRPS daily rainfall data from the year 2020.^50^ If one unit was parameterized for both urban and rural settings, we combined the two by the proportion of the population living in each setting so that there was only one distinct parameter set assigned to each site. Since models were run into the future through to the year 2032, years 2023-2032 were parameterized with 2022 intervention coverage levels assumed to be kept constant over the remainder of the simulation period.

Each metapopulation model run began with the selection of a single seed site, defined as the centroid of an administrative unit with an international border. The three closest neighboring units within the same country as the seed site were also selected (as measured by distance between centroids), as well as the four nearest neighboring sites across an international border from the seed site; the eight total selected sites represented one model run or “cluster” (**Figure 1C**). 401 clusters were formed with seed sites representing each of the 401 administrative units touching an international border. Clusters allow for interpretation of trends in malaria in border regions, but do not account for importation of infection from a border area to population centers further inland.

Mixing matrices were formed for each cluster of eight sites using the gravity model process described above, using a grid comprised of 0.1×0.1 degree cells and summarizing at the first administrative unit level. Origin and destination points were assigned to geo-spatial grid centroids and the travel times between grid centroids were set using an algorithm to calculate the path-of-least-resistance from origin to destination across a pixelated friction surface^14^ created by the Malaria Atlas Project which accounts for land type, and presence of roads or water (**Figure 1D**). The resulting travel times and administrative unit population sizes^41^ (**Figure 1E**) were used to create a mixing matrix for the metapopulation model (**Figure 1F**). Each of the 401 site models were run with 50 random draws from the main model parameter distributions to generate uncertainty estimates.

### Border intervention

Border posts were implemented in the model by modifying the influence of EIR and FOIM between sites by a coefficient ranging between 0 and 1. Coefficients are functions of the estimated percent of travelers captured by border posts (as opposed to travel through un-surveilled routes), RDT sensitivity, and treatment efficacy (**Figure S5**). A value of 0 means that the border post halts all transmission, and a value of 1 indicates that the border post has no effect. A value of 0.2 representing 80% coverage (80% of travelers “screened” at a border post) was used for all site runs. This value represents an optimistic best-case scenario. A sensitivity analysis was performed to vary coverage, but the feasibility of implementing border posts along international borders is likely to be challenging and country specific. RDT sensitivity is prevalence-dependent, meaning that RDTs are less sensitive to detecting infections in low *Pf*PR_2-10_ settings and highly sensitive in high *Pf*PR_2-10_ settings, due to a greater proportion of infections with higher parasitemia levels. RDT sensitivity curves were obtained from previously published parameters (**Figure S6**),^29,30^ with the assumption that the sum of subpatent, infected, and clinically diseased individuals at each time point was the true PCR prevalence. Treatment was assumed to be effective in 95% of treated individuals.^51^ Border posts represent real world interventions such as static border posts at border entry points, or mobile malaria posts along border crossing areas and community focal points frequented by target populations.^13,32^ Border posts were only implemented between sites across an international border; no posts were implemented between sites falling within the same country. All border posts were assumed to capture bi-directional movement across international borders.

### Analysis

The primary outcome of interest was the number of cases per year averted in each unit (accounting for unit population size) after the implementation of border posts. Secondary outcomes included the change in *Pf*PR_2-10_ after the intervention and identification of areas where border posts could have the greatest effect on malaria elimination goals. The period of analysis was ten years following border post implementation.

### Case studies

We used two generic case studies to examine the influence of 1) site *Pf*PR_2-10_ and 2) border post coverage on the effectiveness of border post interventions over 10 years. In the first case study we ran 2-unit models with every combination of *Pf*PR_2-10_ ranging from 5% to 80% in 5% step intervals, and 80% intervention coverage. In the second case study we ran 2-unit models with *Pf*PR_2-10_ values of 10%, 20%, 40%, 60% and 80%, and border post coverage assumptions ranging from 0% to 100% in 5% step intervals. Both case studies were set-up assuming equal population sizes (100,000 people per unit), a 20-year warm-up period, and 5% mixing between units. Each 2-unit model was run using 50 unique random draws from the main model parameter distributions to generate uncertainty estimates.

## Data Availability

The data required to fit duration estimates for mixing matrices can be found in Marshall et al. 2016. Unit parameters for epidemiological data, population size, historical intervention coverage, mosquito vectors, and seasonality were obtained from publicly available resources referenced in the methods section. Full analysis code, including model parameterization, unit parameterization, and mixing algorithms are provided at: https://github.com/htopazian/border_elimination.

## Data availability

The data required to fit duration estimates for mixing matrices can be found in Marshall et al. 2016. Unit parameters for epidemiological data, population size, historical intervention coverage, mosquito vectors, and seasonality were obtained from publicly available resources referenced in the methods section.

## Code availability

Full analysis code, including model parameterization, unit parameterization, and mixing algorithms are provided at: https://github.com/htopazian/border_elimination.

## Acknowledgements

We thank Dr. Kim Lindblade (PATH) for her input on border posts in near-elimination settings.

This work was supported by the Wellcome Trust [reference 220900/Z/20/Z]. HMT, GDC, NS, MP, KH, and ACG acknowledge funding from the MRC Centre for Global Infectious Disease Analysis (reference MR/X020258/1), funded by the UK Medical Research Council (MRC). This UK funded award is carried out in the frame of the Global Health EDCTP3 Joint Undertaking.

KH is additionally funded by the National Institute for Health and Care Research (NIHR) Health Protection Research Unit in Modelling and Health Economics, a partnership between the UK Health Security Agency, Imperial College London and LSHTM [grant code NIHR200908]. KH also acknowledges funding from Community Jameel and by Kenneth C. Griffin. Disclaimer: *“The views expressed are those of the author(s) and not necessarily those of the NIHR, UK Health Security Agency or the Department of Health and Social Care*.*”*

For the purpose of open access, the authors have applied a ‘Creative Commons Attribution’ (CC BY) license to any Author Accepted Manuscript version arising from this submission.

## Author contributions

Study conception and design: HMT, GDC, ACG; built models and collated data: HMT, GDC; carried out analysis: HMT; interpreted data: HMT, GDC, NS, MP, KH, ACG; drafted manuscript: HMT; substantively reviewed drafts and approved manuscript for submission: all authors.

## Additional information

Correspondence and requests for materials should be addressed to Hillary M. Topazian.

