## Supplementary information for "Estimating the potential impact of surveillance test-and-treat posts to reduce malaria in border regions in sub-Saharan Africa: a modelling study"

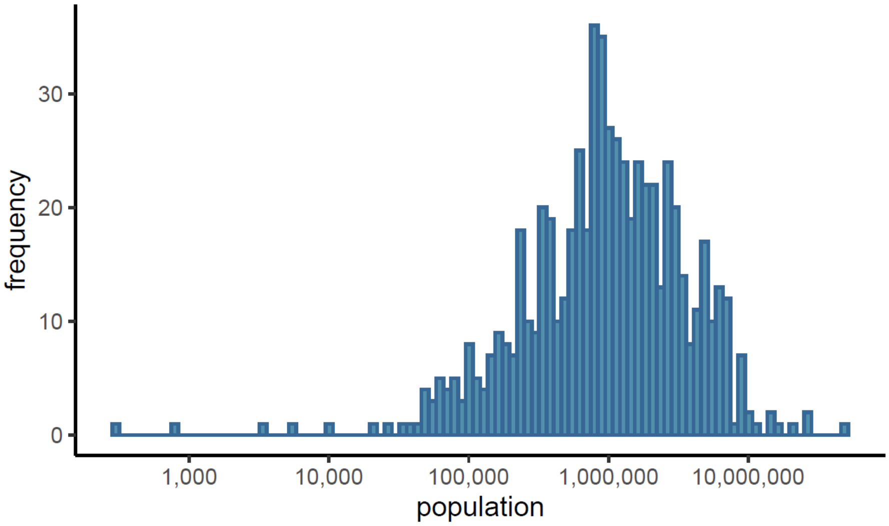


**Figure S1**. **Distribution of administrative unit population sizes**. N = the 636 administrative units identified in malaria-endemic sub-Saharan Africa. Population counts are extracted from WorldPop 2023.[1]

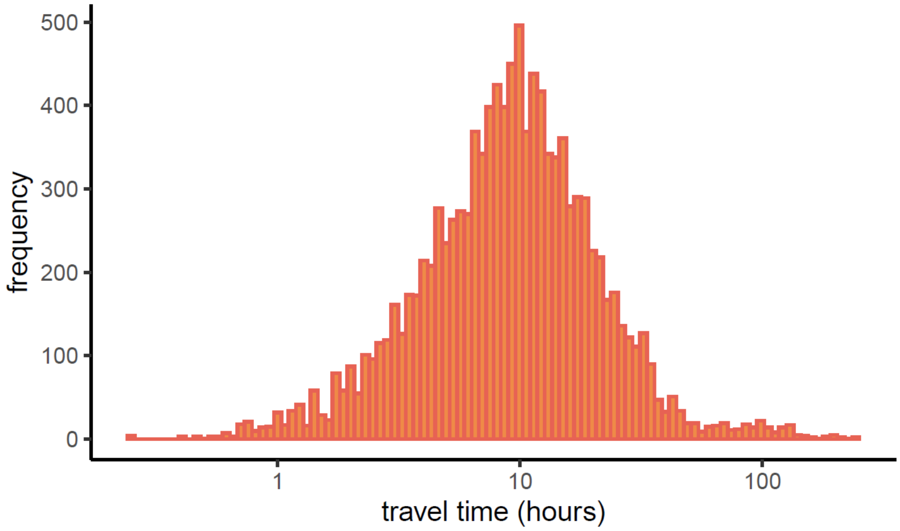


**Figure S2**. **Distribution of travel times (hours)**. Travel times were measured between 8 sites in each of 401 cluster pairs identified using seed points from administrative units in malaria-endemic sub-Saharan Africa which touch international borders. Travel times were estimated using an algorithm to calculate the path-of-least-resistance from origin to destination across a friction surface[2] taken from the Malaria Atlas Project.

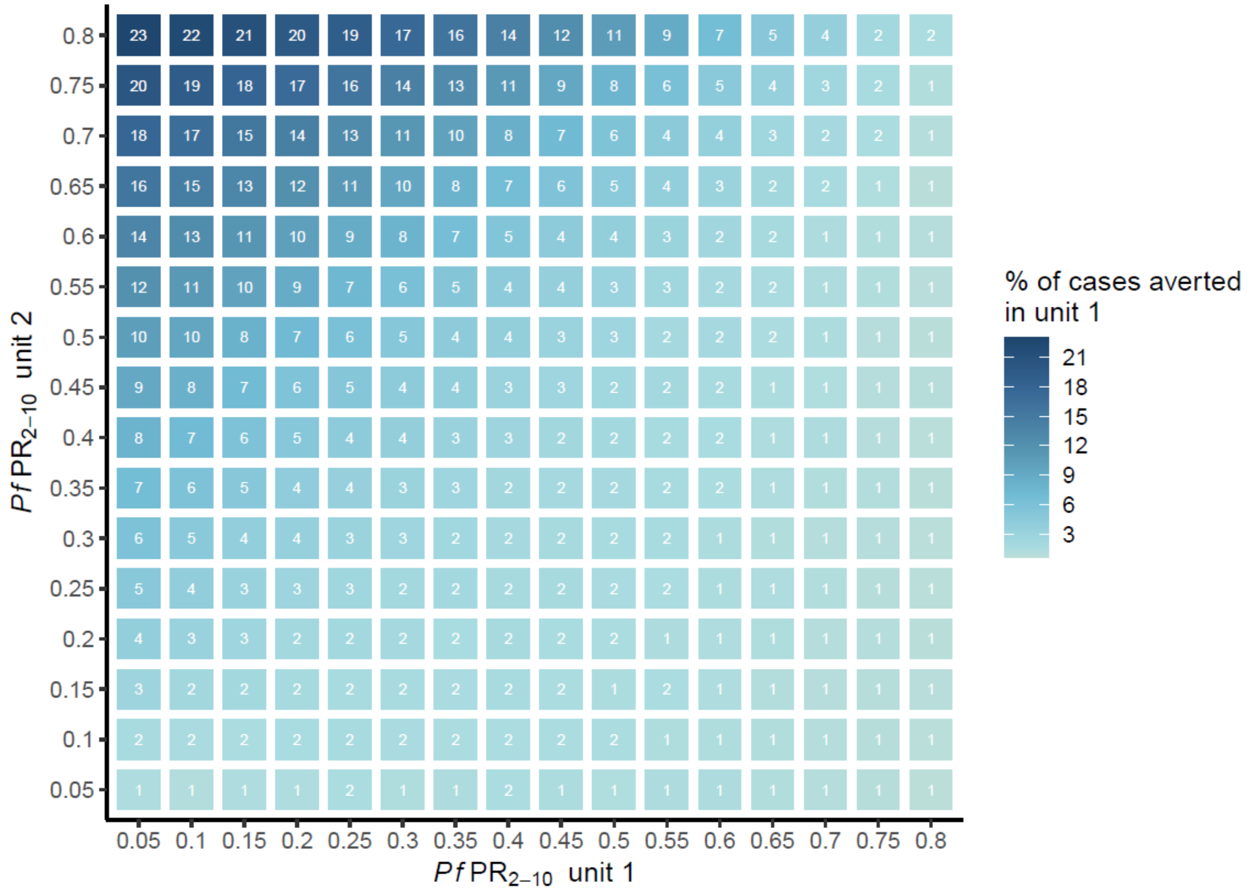


**Figure S3**. **Measuring the sensitivity of border posts to *Pf*PR$_{2-10}$**. Sensitivity analysis to a case-study examining the effects of *Pf*PR$_{2-10}$ on the percent of cases averted using a 2-unit metapopulation model over a 10-year period. Cases are detected via PCR as opposed to the rapid diagnostic tests used in the primary analysis. Colors represent median values out of 50 unique parameter draws. The model assumes that 95% of mixing occurs within a unit and 5% of mixing between units.

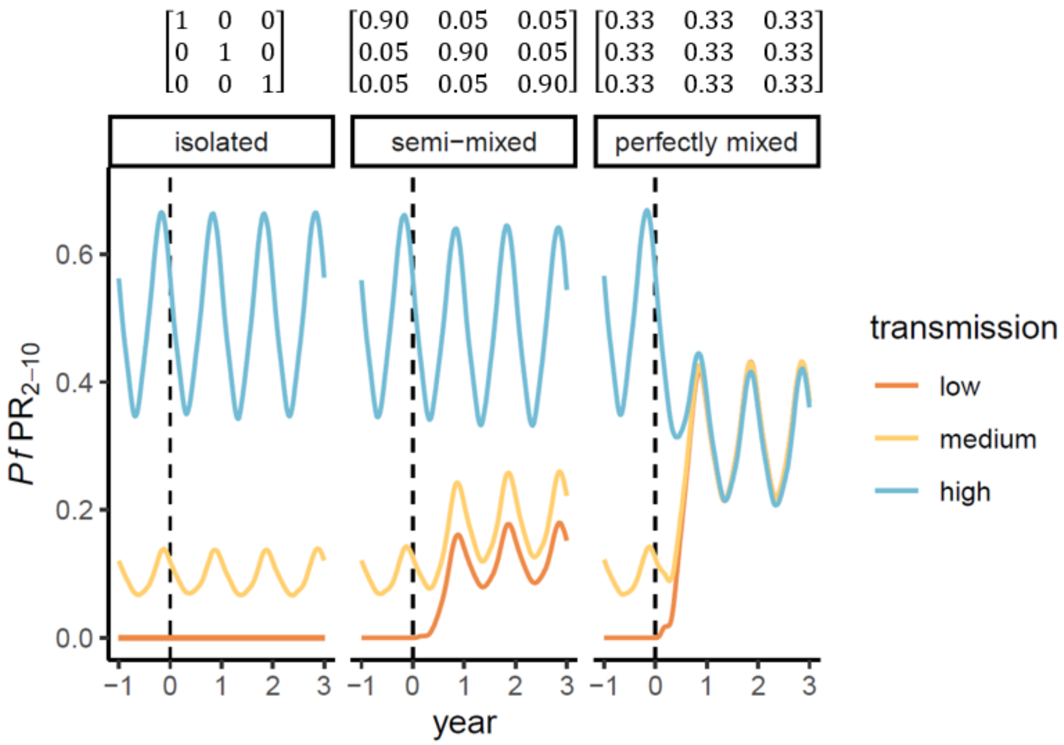


**Figure S4**. **Illustrative 3-unit metapopulation model**. The model plots the effects of different mixing patterns on *Pf*PR$_{2\text{-}10}$ over time. Each unit has a unique level of malaria transmission. Mixing matrices are shown above each panel where rows are equal to the origin site (low, medium, high) and columns are equal to the destination site (low, medium, high). Diagonal matrix values indicate within-unit mixing. The dotted line represents the time at which mixing between units begins.

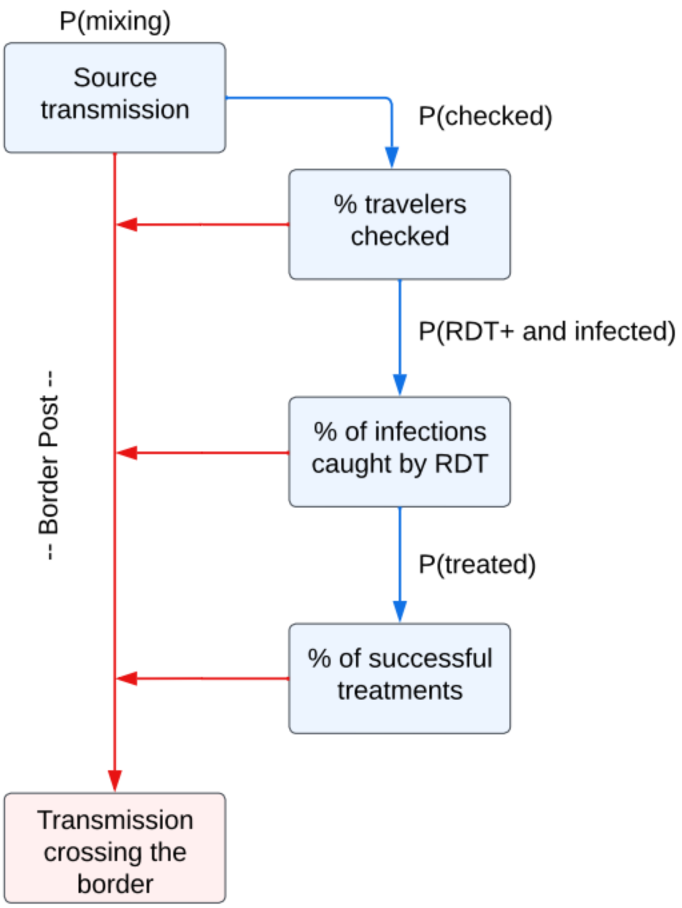


**Figure S5**. **Border post flow diagram**. Flow diagram illustrating the mechanisms by which border posts reduce *P. falciparum* transmission. The model assumes that a border post captures a certain percentage of human mixing (as measured through the entomological inoculation rate and the force of infection on mosquitos), that rapid diagnostic tests are <100% sensitive to detecting true infections,[3,4] and that treatment with artemether-lumefantrine is successful in 95% of treated individuals.[5] Note: RDT = rapid diagnostic test, and RDT+ indicates a positive test.

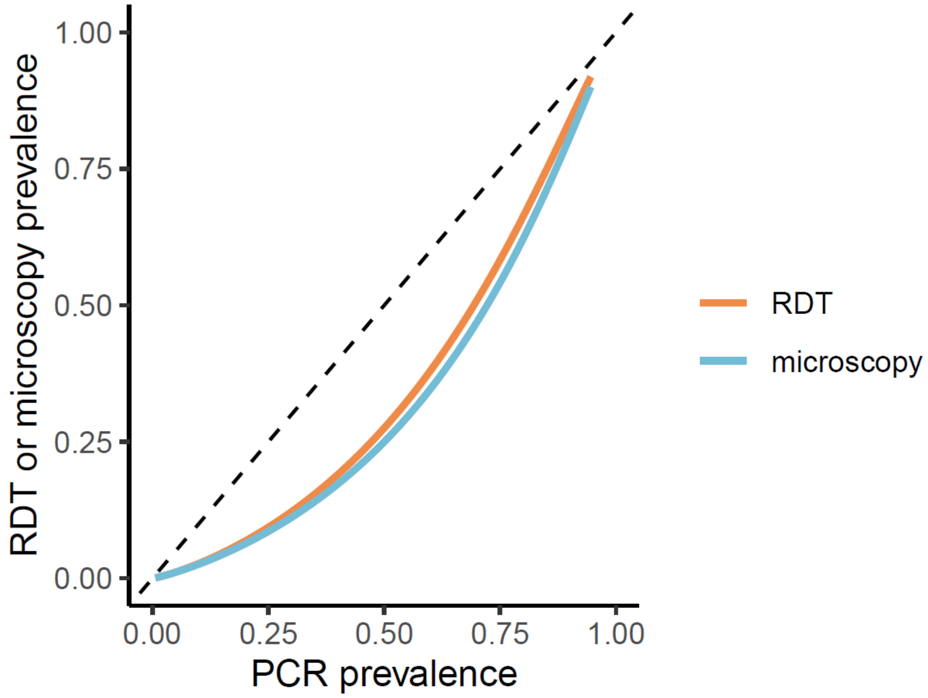


**Figure S6**. **PCR vs. rapid diagnostic tests (RDT) prevalence**. Fits are taken from previously published parameters[3,4] using RDT, microscopy, and PCR data across various geographies and age groups. Note: PCR = polymerase chain reaction, RDT = rapid diagnostic test.

### TECHNICAL METHODS

#### Transmission model

*malariasimulation* (*v1.3.0*; Charles G, et al. 2022) is an open-source individual-based mathematical model of *P. falciparum* malaria. Individuals enter the model at birth as susceptible to *P. falciparum* infection ($S$) (**Figure S7**). Maternal immunity is modelled from birth and exponentially decays over the first 6 months of an individual's life. Throughout the life course, individuals become exposed to infectious mosquito bites. The hazard of becoming infected is dependent on the force of infection from mosquito to human ($\Lambda_i$), which is itself dependent on an individual's level of pre-erythrocytic immunity as well as vector characteristics such as biting rate, infectiousness, and population size. Once infected, individuals pass through a 12-day latent period ($d_E$). Individuals then develop either asymptomatic infection ($A$) or clinical disease ($D$) with a probability ($\phi_i$) which is dependent on an individual's existing level of immunity against clinical disease. A proportion of individuals with clinical disease are treated with probability ($f_T$); treated individuals move to infection state $T$. A proportion of individuals with clinical disease also go on to develop severe disease. Treated individuals recover from infection at rate $r_T$ and acquire a waning prophylactic protection from being reinfected; when this protection disappears, individuals return to the susceptible state ($S$). Individuals not receiving treatment move to diseased state $D$, from $D$ to $A$ at rate $r_D$, then from $A$ to a sub-patent infection state ($U$) at rate $r_A$ (due to declining parasite density), then from $U$ to $S$ at rate $r_U$. Super-infection (re-infection) via another infectious bite can occur for individuals in states $D$, $A$ and $U$.

Deaths in the model are calculated using national life tables, where individuals die and exit the model at age-specific rates which align with national age distributions. All deaths are due to natural causes; the model does not include malaria-related mortality, although deaths from malaria can be estimated after simulations are run as a proportion of those experiencing severe disease (0.215).[6] Across the simulation period, the total population size remains constant as deaths become replaced with births of new individuals entering the model with identical individual biting rates.

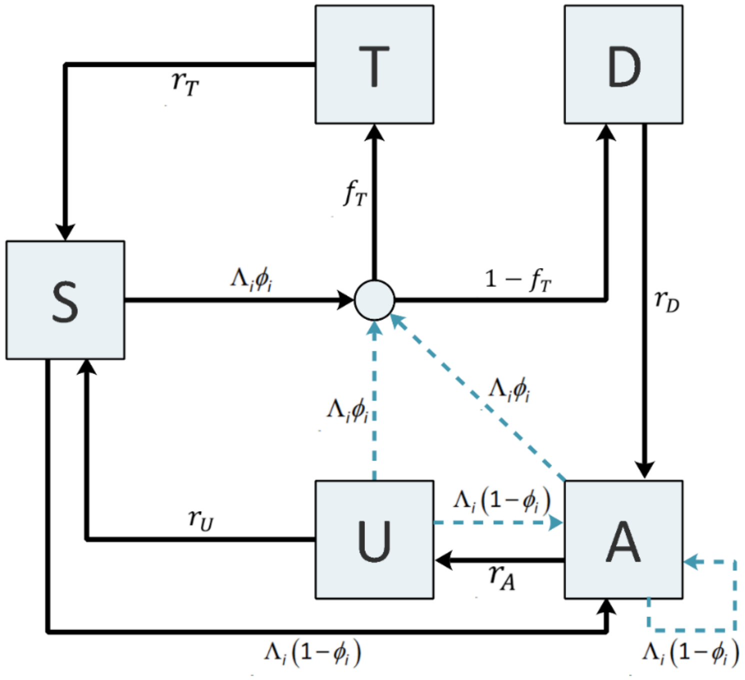


**Figure S7**. **Flow diagram for the human *P. falciparum* transmission model**. This illustration has been previously published by Winskill et al. 2017.[7] States are shown in boxes and transitions are shown via arrows; dashed arrows represent superinfection. Hazard rates are shown next to transition arrows. The circle in the center of the diagram represents the treatment node. Note: S = susceptible; T = treated clinical disease; D = untreated clinical disease; A = asymptomatic patent infection; U = asymptomatic sub-patent infection. All parameters and rates are described and referenced within **Table S1**.

**Table S1. Infection state transition rates between human compartments**.

| **Process** | **Transition** | **Rate** |
|---|---|---|
| Infection | S → I | $\Lambda_i(t-d_E)$ |
| Progression of untreated disease to asymptomatic infection | D → A | $r_D = \frac{1}{d_D}$ |
| Progression of asymptomatic infection to sub-patent infection | A → U | $r_A = \frac{1}{d_A}$ |
| Progression of sub-patent infection to susceptible | U → S | $r_U = \frac{1}{d_U}$ |
| Progression of treated disease to susceptible (treated individuals experience a period of drug-dependent partial protection from reinfection) | T → S | $r_T = \frac{1}{d_T}$ |
| Super-infection from untreated clinical disease, asymptomatic, or sub-patent infection | D → I<br>A → I<br>U → I | $\Lambda_i(t-d_E)$ |

#### Mosquito biting rates

Mosquito biting rates are dependent on a variety of factors including heterogeneities in an individual's assigned biting rate at birth, and an age-dependent biting rate due to increasing risk of exposure with increasing body surface area. Age-dependency in biting exposure is modelled through a distribution of the entomological inoculation rate (EIR), the number of infectious bites per individual per timestep, by age. Each individual's unique biting rate is a product of $\zeta_i$, their relative biting rate assigned at birth, and $\psi_i$, their relative age-dependent biting rate for a given age $a$:

$$\psi_i(a) = 1 - \rho exp\left(-\frac{a}{a_0}\right)$$

Here, $\rho$ and $a_0$ determine the relationship between age (via body surface area) and biting rate.

The human adult EIR is given by $\varepsilon = \frac{\alpha I_M}{\omega}$ where $\alpha$ is the rate at which a mosquito feeds, the parameter $\omega$ is a normalizing constant, and $g(a)$ is the cross-sectional human population age-distribution:

$$\omega = \int_0^\infty \psi(a) g(a) da$$

The relative individual biting rate is drawn from a log-normal distribution with a mean of 1, where $\sigma^2$ is the variance of the log heterogeneity in biting rates. This biting rate remains constant over an individual's lifetime:

$$log(\zeta_i) \sim N\left(\frac{-\sigma^2}{2}, \sigma^2\right)$$

The EIR $\varepsilon_i(a,t)$ and force of infection $\Lambda_i(a,t)$ are combinations of individual and the age-specific biting rates, here given for individual $i$ with age $a$ at time $t$:

$$\varepsilon_i(a,t) = \varepsilon_0(t)\, \zeta_i \psi_i(a)$$

$$\Lambda_i(a,t) = b_i(t)\, \varepsilon_i(a,t)$$

$\varepsilon_0(t)$ is the mean EIR experienced by adults at time $t$. $b_i(t)$ is the probability that an infectious bite leads to a patent infection and is dependent on the level of pre-erythrocytic immunity and considers a lag of $d_E$ days to account for the latent period of sporozoite infection after an infectious bite.

#### Immunity

Immunity in the model is captured through several layers corresponding to distinct stages of infection:

1. **Maternal immunity** against clinical disease $I_{CM}$ and severe disease $I_{VM}$ is acquired at birth (placental antibody transfer). Maternal immunity decays exponentially at rate $r_M$ and causes a reduction in the probability of clinical or severe disease following infection. The level of immunity at birth is a proportion $P_M$ of the immunity levels in a representative population aged

15-35 years with the same biting heterogeneity level as the targeted individual. Maternal immunity decays exponentially at rate $r_M$:

$$r_M = \frac{1}{d_M}$$

2. **Pre-erythrocytic immunity** $I_B$ develops in individuals as they age and is boosted each time an individual experiences an infectious bite, provided it has been at least $u_B$ days since the previous exposure. $I_B$ reduces the probability of infection given an infectious bite and decays exponentially after an exposure at rate $r_B$:

$$r_B = \frac{1}{d_B}$$

3. **Blood stage immunity** against severe disease $I_{VA}$ and clinical disease $I_{CA}$ reduces parasite blood densities, leading to reduced probabilities of clinical and severe disease. Blood stage immunity parameters decay exponentially between exposures at rates $r_{VA}$ and $r_C$ for severe and clinical disease, respectively:

$$r_{VA} = \frac{1}{d_{VA}}, r_C = \frac{1}{d_{CA}}$$

Each parameter is boosted following patent infection if it has been at least $u_V$ or $u_C$ days, respectively since the previous exposure. Immunity levels are converted to individual time-dependent probabilities of developing an infection using Hill functions. The probability of developing infection after an infectious bite is represented by the following for individual $i$ at time $t$:

$$b_i(t) = b_0\left(b_1 + \frac{1 - b_1}{1 + \left(\frac{I_B(i,t)}{I_{B0}}\right)^{\kappa_B}}\right)$$

Here $b_0$ represents the probability of infection with no immunity, $b_1$ is the minimum probability of infection, $I_{B0}$ and $\kappa_B$ are shape and scale parameters respectively, and $I_B(i,t)$ is the level of pre-erythrocytic immunity for that specific individual $i$ at time $t$.

The probability of developing clinical disease (given infection) for individual $i$ at time $t$ is given as:

$$\phi_i(\mathrm{t}) = \phi_0\left(\phi_1 + \frac{1 - \phi_1}{1 + \left(\frac{I_{CA}(\mathrm{i},\mathrm{t}) + I_{CM}(i,t)}{I_{C0}}\right)^{\kappa_C}}\right)$$

$\phi_0$ represents the probability of disease with no immunity, $\phi_1$ is the minimum probability of developing clinical disease, $I_{C0}$ and $\kappa_C$ are scale and shape parameters respectively, $I_{CA}(i,t)$ is the level of acquired immunity to clinical disease, and $I_{CM}(i,t)$ is the level of maternally acquired immunity to clinical disease.

The probability of developing severe disease (given infection) is represented for individual $i$ at time $t$ is given as:

$$\theta_i(a,t) = \theta_0\left(\theta_1 + \frac{1-\theta_1}{1+f_V(i,a)\left(\frac{(I_{VA}(i,t)+I_{VM}(i,t)}{I_{V0}}\right)^{\kappa_V}}\right)$$

$\theta_0$ represents the probability of severe disease with no immunity, $\theta_1$ is the minimum probability, $I_{V0}$ and $\kappa_V$ are scale and shape parameters respectively, $I_{VA}(i,t)$ is the level of acquired immunity to severe disease, and $I_{VM}(i,t)$ is the level of maternally acquired immunity to severe disease. $f_V(i,a)$ is an age-dependent modifier of the risk of severe disease with $f_{V0}, a_V$ and $\gamma_V$ parameters:

$$f_V(i,a) = 1 - \frac{(1-f_{V0})}{\left(1+\left(\frac{a}{a_V}\right)^{\gamma_V}\right)}$$

Treated individuals have a reduced probability of moving from clinical disease to severe disease, $f_{VT}$.

4. **Detection immunity** (blood stage) against detectability of asymptomatic infection $I_D$ reduces both the probability of detection by microscopy and infectiousness to mosquitoes and decays exponentially at rate $r_{ID}$:

$$r_{ID} = \frac{1}{d_{ID}}$$

$r_{ID}$ is boosted following each patent infection provided it has been at least $u_D$ days since the previous exposure.

The detectability of an asymptomatic infection by microscopy in an individual $i$ of age $a$ at time $t$ is represented by:

$$q_i(a,t) = d_1 + \frac{1-d_{min}}{\left(\left(\frac{1+I_D(i,t)}{I_{D0}}\right)^{\kappa_D} f_D(i,a)\right)}$$

Here $d_{min}$ is the minimum probability of detection, $I_{D0}$ and $\kappa_D$ are scale and shape parameters respectively, $I_D(i,t)$ is the level of acquired immunity to the detectability of infection of individual $i$ at time $t$ and $f_D(i,a)$ is an age-dependent modifier of the risk of detectability with $f_{D0}, a_D$ and $\gamma_D$ parameters:

$$f_D(i,a) = 1 - \frac{(1-f_{D0})}{\left(1+\left(\frac{a}{a_D}\right)^{\gamma_D}\right)}$$

Immunity to detection causes a reduction in parasite density which lowers the probability of transmission. Onward transmission is possible in all infectious states, but highest infectivity is associated with diseased states. In states $D$ and $U$, onwards infectiousness is represented by $c_D$ and $c_U$ respectively, and $c_T$ after treatment. In state $A$, infectiousness is modified by $q_i$, the detectability of infection from individual $i$: $c_A = c_U + (c_D + c_U)\, q_i^{\gamma_1}$.

#### Vector Model

The vector model is based on a compartmental stochastic structure[8] of the life cycle of female *Anopheles spp.* mosquitoes (which transmit *P. falciparum* parasites). *An. gambiae s.s*, *An. arabiensis*, and *An. funestus* are the three species captured by the model and are all parameterized with unique bionomics (**Table S2**).

Adult female mosquitos lay eggs at rate $\beta$. Hatched larvae progress through early ($E$) and late larval stages ($L$) and into the pupal developmental stage ($P$). The larval stages are regulated by a density dependent mortality based on a time-varying carrying capacity, $K$, representing the ability of the environment to sustain breeding sites through the year. The parameter $\gamma$ regulates the larval density relative to the carrying capacity, and ultimately determines the adult mosquito density and transmission intensity in scenarios without interventions. It is assumed that 50% of emerging adult mosquitoes are female and enter the susceptible state $S_M$. Adult female mosquitoes become infected at rate $\Lambda_M$, a function of the infectiousness of humans. $\Lambda_M$ contains a time-lag, $t_l$, to account for the incubation time between when a human becomes infected and subsequently becomes infectious. Similarly, there is a fixed delay $\tau_M$ before an infected female mosquito subsequently becomes infectious to humans ($I_M$). It is assumed that infected adult mosquitoes remain infectious until death.

**Table S2**. **Vector bionomics parameters, stratified by *Anopheles* species**.

| Bionomics trait | *An. gambiae s.s* | *An. arabiensis* | *An. funestus* |
|---|---|---|---|
| Anthropophagy | 0.92 | 0.71 | 0.94 |
| % bites indoors | 0.97 | 0.96 | 0.98 |
| % bites indoors and in bed | 0.89 | 0.90 | 0.90 |
| Daily mortality of adults with no interventions | 0.132 | 0.132 | 0.112 |

#### Seasonality

Seasonality is included in the model through modifications of the environment's carrying capacity of mosquitos, $K$, at time $t$:

$$K(t) = K_0 \frac{R(t)}{\bar{R}}$$

$K_0$ is the baseline carrying capacity, $\bar{R}$ is the mean rainfall over a year-long period, and $R(t)$ is the time-varying seasonal curve of interest, obtained from rainfall data using the first three frequencies of a Fourier transformation where $g$ and $h$ are fitted parameters. Daily rainfall data were pulled from the Climate Hazards Group InfraRed Precipitation with Station data (CHIRPS).[9]

$$R(t) = g_0 + \sum_{i=1}^{3} g_i \, cos(\, 2\pi t i) + h_i \, sin(\, 2\pi t i)$$

**Model Parameter Values**

**Table S3.** ***P. falciparum*** **mosquito model and intervention parameter values**. Full details can be found in the original publications[10,11] including references for parameters and intervals for the prior and posterior distributions (median values of the posterior distribution are used in model simulations).

| Parameter | ***malariasimulation*** **parameter** | Definition | Value |
|---|---|---|---|
| **Fixed state transitions** | | | |
| $d_D = \frac{1}{r_D}$ | **dd** | the delay for humans to move from state D to A | 5 |
| $d_T = \frac{1}{r_T}$ | **dt** | the delay for humans to move from state Tr to Ph | 5 |
| $d_A = \frac{1}{r_A}$ | **da** | the delay for humans to move from state A to U | 200 |
| $d_U = \frac{1}{r_U}$ | **du** | the delay for humans to move from state U to S | 110 |
| $d_E$ | **del** | the delay for mosquitos to move from state E to L | 6.64 |
| $d_L$ | **dl** | the delay for mosquitos to move from state L to P | 3.72 |
| $d_P$ | **dpl** | the delay mosquitos to move from state P to $S_m$ | 0.643 |
| **Biting rate** | | | |
| $\sigma^2$ | **sigma_squared** | heterogeneity parameter | 1.67 |
| | **n_heterogeneity_groups** | number of discrete groups for heterogeneity in exposure to mosquito bites | 5 |
| $a_0$ | **a0** | age dependent biting parameter | 2920 |
| $\rho$ | **rho** | age dependent biting parameter | 0.85 |
| **Immunity decay** | | | |

| | | | |
|---|---|---|---|
| $d_M$ | **rm** | decay rate for maternal immunity to clinical disease | 67.6952 |
| $d_{VM}$ | **rvm** | decay rate for maternal immunity to severe disease | 76.8365 |
| $d_B$ | **rb** | decay rate for acquired pre-erythrocytic immunity | 3650 |
| $d_{CA}$ | **rc** | decay rate for acquired immunity to clinical disease | 10950 |
| $d_{VA}$ | **rva** | decay rate for acquired immunity to severe disease | 10950 |
| $d_{ID}$ | **rid** | decay rate for acquired immunity to detectability | 3650 |
| **Pre-erythrocytic infection** | | | |
| $b_0$ | **b0** | maximum probability due to no immunity | 0.59 |
| $b_1$ | **b1** | maximum reduction due to immunity | 0.5 |
| $I_{B0}$ | **ib0** | scale parameter | 43.9 |
| $\kappa_B$ | **kb** | shape parameter | 2.16 |
| **Immunity boost grace periods** | | | |
| $u_B$ | **ub** | period in which pre-erythrocytic immunity cannot be boosted | 7.2 |
| $u_C$ | **uc** | period in which clinical immunity cannot be boosted | 6.06 |
| $u_V$ | **uv** | period in which severe immunity cannot be boosted | 11.4321 |
| $u_D$ | **ud** | period in which immunity to detectability cannot be boosted | 9.44512 |
| **Infectivity towards mosquitos** | | | |

| | | | |
|---|---|---|---|
| $c_D$ | **cd** | infectivity of clinically diseased humans towards mosquitos | 0.068 |
| $\gamma_1$ | **gamma1** | parameter for infectivity of asymptomatic humans | 1.82425 |
| $c_U$ | **cu** | infectivity of sub-patent infection | 0.0062 |
| $c_T$ | **ct** | infectivity of treated infection | 0.021896 |
| **Clinical infection** | | | |
| $\phi_0$ | **phi0** | maximum probability due to no immunity | 0.792 |
| $\phi_1$ | **phi1** | maximum reduction due to immunity | 0.00074 |
| $I_{C0}$ | **ic0** | scale parameter | 18.02366 |
| $\kappa_C$ | **kc** | shape parameter | 2.36949 |
| **Severe disease** | | | |
| $\theta_0$ | **theta0** | maximum probability due to no immunity | 0.0749886 |
| $\theta_1$ | **theta1** | maximum reduction due to immunity | 0.0001191 |
| $\kappa_V$ | **kv** | shape parameter | 2.00048 |
| $f_{V0}$ | **fv0** | age dependent modifier | 0.141195 |
| $a_V$ | **av** | age dependent modifier | 2493.41 |
| $\gamma_V$ | **gammav** | age dependent modifier | 2.91282 |
| $I_{V0}$ | **iv0** | scale parameter | 1.09629 |

| | | | |
|---|---|---|---|
| **Incubation periods** | | | |
| $d_E$ | **de** | duration of the human latent period of infection | 12 |
| $d_g$ | **delay_gam** | lag from parasites to infectious gametocytes | 12.5 |
| $d_{EM}$ | **dem** | extrinsic incubation period in mosquito population model | 10 |
| **Immunity reducing probability of infection** | | | |
| $f_{D0}$ | **fd0** | time-scale at which immunity changes with age | 0.007055 |
| $a_D$ | **ad** | scale parameter relating age to immunity | 7993.5 |
| $\gamma_D$ | **gammad** | shape parameter relating age to immunity | 4.8183 |
| $d_1$ | **d1** | minimum probability due to immunity | 0.160527 |
| $I_{D0}$ | **id0** | scale parameter | 1.577533 |
| $\kappa_D$ | **kd** | shape parameter | 0.476614 |
| **Mortality parameters** | | | |
| $P_{CM}$ | **pcm** | new-born clinical immunity relative to mother's | 0.774368 |
| $P_{VM}$ | **pvm** | new-born severe immunity relative to mother's | 0.195768 |
| **Seasonality** | | | |
| $g_0$ | **g0** | rainfall Fourier parameter | Site-specific |
| $g$ | **g** | rainfall Fourier parameter | |

| | | | |
|---|---|---|---|
| $h$ | **h** | rainfall Fourier parameters | |
| $\gamma$ | **gamma** | effect of density dependence on late instars relative to early instars | 13.25 |
| | **rainfall_floor** | the minimum rainfall value | 0.001 |
| **Mortality parameters** | | | |
| $\mu_E$ | **me** | early stage larval mortality rate | 0.0338 |
| $\mu_L$ | **ml** | late stage larval mortality rate | 0.0348 |
| $\mu_P$ | **mup** | the rate at which pupal mosquitos die | 0.249 |
| $\mu_M$ | **mum** | the rate at which developed mosquitos die | 0.1253333 |
| **Vector biology** | | | |
| $\beta$ | **beta** | the average number of eggs laid per female mosquito per day | 21.2 |
| **1 /** $\delta$ | **blood_meal_rates** | the blood meal rates for each species | 0.3333333333 |
| $Q_0$ | **Q0** | proportion of blood meals taken on humans | *A. arabiensis*: 0.71<br>*A. funestus*: 0.94<br>*A. gambiae*: 0.92 |
| | **foraging_time** | time spent taking blood meals | 0.69 |
| **Insecticide-treated nets** [12] | | | |
| $r_N$ | **rn** | probability mosquito is repelled by the bednet | Site-specific |
| $r_{NM}$ | **rnm** | minimum probability mosquito is repelled by the bednet | 0.24 |
| $d_{N0}$ | **dn0** | probability killed by the bednet | Site-specific |

| | | | |
|---|---|---|---|
| $\Phi_B$ | **phi_bednets** | proportion of bites taken in bed | *A. arabiensis*: 0.9<br>*A. funestus*: 0.9<br>*A. gambiae*: 0.89 |
| $\Phi_I$ | **phi_indoors** | proportion of bites taken indoors | *A. arabiensis*: 0.96<br>*A. funestus*: 0.98<br>*A. gambiae*: 0.97 |
| $k_0$ | **k0** | proportion of female mosquitoes bloodfed with no net | 0.699 |
| | **gamman** | the half-life of bednet efficacy (timesteps) | Site-specific |
| **Indoor residual spraying** [13] | | | |
| $l_{s\vartheta}$ | **ls_theta** | maximum killing effect for a given IRS | Site-specific |
| $l_{s\gamma}$ | **ls_gamma** | how quickly IRS killing effect wanes | Site-specific |
| $k_{s\vartheta}$ | **ks_theta** | maximum blood-feeding inhibition effect for a given IRS | Site-specific |
| $k_{s\gamma}$ | **ks_gamma** | how quickly IRS blood-feeding inhibition effect wanes | Site-specific |
| $m_{s\vartheta}$ | **ms_theta** | maximum deterrence effect for a given IRS | Site-specific |
| $m_{s\gamma}$ | **ms_gamma** | determines how quickly IRS deterrence effect wanes | Site-specific |
| **Treatment** | | | |
| $P_T$ | **drug_efficacy** | a vector of efficacies for available drugs | AL: 0.95<br>SP: 0.90<br>SP + AQ: 0.90 |
| | **drug_rel_c** | a vector of relative onwards infectiousness values for drugs | AL: 0.0509<br>SP: 0.3200<br>SP + AQ: 0.3200 |
| | **drug_prophilaxis_shape** | a vector of shape parameters for weibull curves to model prophylaxis for each drug | AL: 11.3<br>SP: 2.516<br>SP + AQ: 3.40 |
| | **drug_prophilaxis_scale** | a vector of scale parameters for weibull curves to model prophylaxis for each drug | AL: 10.6<br>SP: 46.68<br>SP + AQ: 39.34 |
| $f_T$ | **clinical_treatment_coverages** | a vector of coverage values for each drug | Site-specific |

| | | | |
|---|---|---|---|
| **RTS,S** | | | |
| $\boldsymbol{V_{max}}$ | **rtss_vmax** | the maximum efficacy of the vaccine | 0.93 |
| $\boldsymbol{\alpha}$ | **rtss_alpha** | shape parameter for the vaccine efficacy model | 0.74 |
| $\boldsymbol{\beta}$ | **rtss_beta** | scale parameter for the vaccine efficacy model | 99.4 |
| $\boldsymbol{CSP_{peak}}$ | **rtss_cs** | peak parameters for the antibody model (mean and std. dev) | 6.37008, 0.35 |
| $\boldsymbol{CSP_{boost}}$ | **rtss_cs_boost** | peak parameters for the antibody model for booster rounds (mean and std. dev) | 5.56277, 0.35 |
| $\boldsymbol{\rho_{peak}}$ | **rtss_rho** | delay parameters for the antibody model (mean and std. dev) | 2.37832, 1.00813 |
| $\boldsymbol{\rho_{boost}}$ | **rtss_rho_boost** | delay parameters for the antibody model for booster rounds (mean and std. dev) | 1.03431, 1.02735 |
| $\boldsymbol{d_s}$ | **rtss_ds** | delay parameters for the antibody model, short-term weaning (mean and std. dev) | 3.74502, 0.341185 |
| $\boldsymbol{d_l}$ | **rtss_dl** | delay parameters for the antibody model, long-term weaning (mean and std. dev) | 6.30365, 0.396515 |
| **Demography** | | | |
| | **deathrate_agegroups** | vector of agegroups (days) | Site-specific |
| | **deathrates** | deathrates per age group | Site-specific |

**Intervention Models**

A range of anti-malaria interventions are supported by the model:

Treatment

Treated individuals move from a clinically diseased state to a susceptible state, retaining a drug-dependent period of prophylaxis against re-infection. Two types of drugs are used in the analysis, artemether-lumefantrine (AL) and sulphadoxine-pyrimethamine (SP). AL has a 95% probability of successfully clearing infection and parameters were originally estimated using a pharmacokinetic-pharmacodynamic model fit to clinical trial data from six sites across sub-Saharan Africa.[5]

Protection from infection at time $u$ after effective treatment is represented by $P_T(u)$ and the probability of re-infection is multiplied by $1 - P_T(u)$. The overall level of protection can be calculated by taking the area under the curve:

$$A_T = \int_0^\infty P_T(u)\ du$$

Treatment results in an overall level of protection, $A_T$, which varies from 7 to 16 days (depending on age) under treatment with AL and 25 days under treatment with SP.

Insecticide-treated bed nets and indoor residual spraying

Insecticide-treated bed nets (ITNs) and indoor residual spraying (IRS) are modelled as malaria prevention tools which either repel female mosquitoes from biting a human host or kill mosquitoes as they attempt to feed. There are six outcomes of an attempted mosquito feed, which are modelled probabilistically:

1. The mosquito bites a non-human host.
2. The mosquito is killed by the ITN before biting.
3. The mosquito is killed by IRS before biting.
4. The mosquito is killed by IRS after biting.
5. The mosquito successfully feeds and survives.
6. The mosquito is repelled by the ITN or IRS without biting and will attempt to find an alternative blood meal source.

The probability of a successful mosquito feed is dependent on the vector species, existing interventions, and level of insecticide resistance. It is assumed that humans reside in the house, livestock reside outside the house, and that mosquitoes which enter indoors will attempt to bite humans.

The probability that a human host $i$ is bitten during a single feeding attempt is represented by $y_i$. The probability that a mosquito bites a host and survives is represented by $w_i$, and the probability that a mosquito is repelled by ITNs or IRS without biting is represented by $z_i$. Natural vector mortality is not modelled. For an individual with no ITN or IRS protection, $y_i = w_i = 1$ and $z_i = 0$.

The probability that a female mosquito successfully feeds during a single attempt on a human or animal host is represented by:

$$W = (1 - Q_0) + Q_0 \sum_i \pi_i w_i$$

and the probability that a female mosquito is repelled without feeding has probability:

$$Z = Q_0 \sum_i \pi_i z_i$$

$Q_0$ is the proportion of bites taken on humans in settings where interventions are absent and $\pi_i$ is the proportion of bites that individual $i$ receives in settings without interventions.

The mosquito feeding rate $f_R$ is:

$$f_R = \frac{1}{\delta_1 + \delta_2}$$

Where $\delta_1$ and $\delta_2$ represent the time durations in which a mosquito looks for a blood meal and rests between feeds, respectively. $\delta_2$ is not affected by interventions, while $\delta_1$ is represented by $\delta_1 = \frac{\delta_{10}}{1-Z}$ when interventions are in place, where $\delta_{10}$ is the duration of time a mosquito looks for a blood meal in the absence of interventions.

In settings where interventions are absent, $p_1$ and $p_2$ represent the probability that a mosquito survives periods of feeding and resting where $\mu_0$ is the natural mosquito death rate:

$$p_{10} = \exp(-\mu_0 \delta_{10})$$

$$p_2 = \exp(-\mu_0 \delta_2)$$

Under the presence of interventions, $p_2$ remains as is, and $p_1$ becomes:

$$p_1 = \frac{p_{10}\, W}{1 - Z\, p_{10}}$$

The probability of a mosquito surviving a single feeding cycle is calculated by multiplying $p_1$ and $p_2$. The mosquito death rate ($\mu_M$) then becomes:

$$p_1 p_2 = \exp\left(-\frac{\mu}{f_R}\right)$$

$$\mu = -f_R \log(p_1 p_2)$$

The probability that a mosquito survives through the extrinsic incubation period ($p_M$) changes as $\mu$ changes.

The probability that a single feeding attempt $q$ ends with a successful bite on human $i$ is given by:

$$q_i = p_{10}(Q_0 \pi_i w_i + Z\, q_i)$$

$$q_i = \frac{p_{10} Q_0 \pi_i w_i}{1 - Z\, p_{10}}$$

The probability that a feeding attempt ends with a bite on an animal host, $q_A$, is:

$$q_A = p_{10}(1 - Q_{10} + Z\, q_A)$$

$$q_A = \frac{p_{10}(1 - Q_0)}{1 - Z\, p_{10}}$$

The proportion of successful bites on humans $(Q)$ is given by:

$$Q = 1 - \frac{q_A}{q_A + \sum_i q_i} = 1 - \frac{(1 - Q_0)}{(1 - Q_0) + Q_0 \sum_i \pi_i w_i} = 1 - \frac{(1 - Q_0)}{W}$$

The human biting rate is given by:

$$\alpha = Q\, f_R$$

The rate at which person $i$ is bitten by a given mosquito species is given by:

$$\lambda_i = \pi_i w_i \frac{\alpha}{\sum_i \pi_i w_i}$$

Upon IRS implementation, it is possible that some mosquitoes may successfully bite and subsequently die while resting on a sprayed surface. When this occurs, the biting rate of each individual is modified by $\frac{y_i}{w_i}$, causing the biting rate to become:

$$\tilde{\lambda}_i = \frac{\alpha\, \pi_i\, y_i}{\sum_i \pi_i w_i}$$

The impact of ITNs and IRS is dependent on the proportion of bites a human receives while sleeping under an ITN[6,10] and/or being inside a sprayed house,[6,10] and protection depends on human host movement and sleeping patterns, mosquito behavior, and the intrinsic efficacy of interventions. It is assumed that human movement and sleeping patterns are not dependent on age or relative exposure (due to a lack of data).

Here, $\lambda_I(t)$ is the rate at which a person indoors at time $t$ is bitten, and $\lambda_O(t)$ is the rate at which a person outdoors at time $t$ is bitten. Estimating the proportion of human hosts who are indoors $p_I(t)$ or in bed $p_B(t)$ at time $t$ allows for calculation of the proportion of bites taken while a human host is indoors $(\Phi_I)$ and in bed $(\Phi_B)$:

$$\Phi_I = \frac{\sum_t p_I(t)\lambda_I(t)}{\sum_t \Big(\big(1 - p_I(t)\big)\lambda_O(t) + p_I(t)\lambda_I(t)\Big)}$$

$$\Phi_B = \frac{\sum_t p_B(t)\lambda_I(t)}{\sum_t \Big(\big(1 - p_I(t)\big)\lambda_O(t) + p_I(t)\lambda_I(t)\Big)}$$

The probabilities of a mosquito being repelled, feeding successfully, or dying in the presence of ITNs and/or IRS are given below in **Table S4**, and the parameter values used in the model are included in **Table S3**.

**Table S4**. **Vector control probabilistic model showing outcome probabilities under ITN and/or IRS interventions**.

| Outcome | Probability |
|---|---|
| **Any biting ($y_i$)** | |
| IRS only | $1 - \Phi_I + \Phi_I(1 - r_s)$ |
| ITN only | $1 - \Phi_B + \Phi_B s_N$ |
| IRS and IRN | $1 - \Phi_I + \Phi_B(1 - r_s)s_N + (\Phi_I - \Phi_B)(1 - r_s)$ |
| **Bites and survives ($w_i$)** | |
| IRS only | $1 - \Phi_I + \Phi_I(1 - r_s)\, s_s$ |
| ITN only | $1 - \Phi_B + \Phi_B s_N$ |
| IRS and IRN | $1 - \Phi_I + \Phi_B(1 - r_s)s_N s_s + (\Phi_I - \Phi_B)(1 - r_s)\, s_s$ |
| **Repelled without feeding ($z_i$)** | |
| IRS only | $\Phi_I r_s$ |
| ITN only | $\Phi_B r_N$ |
| IRS and IRN | $\Phi_B(1 - r_s)r_N + \Phi_I r_s$ |

$\boldsymbol{\Phi_I}, \boldsymbol{\Phi_B}$ time dependent probability of feeding indoors and on someone in bed, respectively
$\boldsymbol{r_N}, \boldsymbol{r_s}$ probability of being repelled before entering the house due to ITNs and IRS, respectively
$\boldsymbol{s_N}, \boldsymbol{s_s}$ probability of successfully feeding after entering the house due to ITNs and IRS, respectively

Briefly, the repellency and mortality effects of IRS and ITNs on mosquitoes begin at values $r_{s0}$, $r_{N0}, d_{s0}, d_{N0}$, respectively and then decrease over time. $r_{s0}$ and $d_{s0}$ (repellency and mortality effects of IRS) decrease at a constant rate over time, $\gamma_s$.

$r_N$, the repelling effect of ITNs, decreases from a maximum, $r_{N0}$, to a non-zero level $r_{NM}$, which represents the level of protection from an ITN that no longer has any insecticidal effect (and potentially physical defects such as holes). $d_N$, the killing effect of ITNs, decreases from $d_{N0}$ at a constant rate $\gamma_N$. After ITNs have been distributed, the following equations represent the probabilities of being repelled, dying, and surviving at time *t*:

$$r_N = (r_{N0} - r_{NM})\exp(-t\gamma_N) + r_{NM}$$

$$d_N = d_{N0}\exp(-t\gamma_N)$$

$$s_N = 1 - r_N - d_N$$

IRS effectiveness depends on the degree of endophily, $\chi$ (the proportion of mosquitoes resting on the wall long enough to be killed by the insecticide), which varies between mosquito species. After IRS has been sprayed, the following equations represent the probabilities of being repelled, dying, and surviving at time *t*:

$$r_s = r_{s0}\exp(-t\gamma_s)$$

$$d_s = \chi d_{s0}\exp(-t\gamma_s)$$

$$s_s = 1 - d_s$$

User inputs for IRS in the model are not parameterized simply by $r_s$, $d_s$, and $s_s$, but by initial effects and waning effects of the intervention dependent on species and type of insecticide. The proportion of mosquitoes dying after entering the indoors, $l_s$, is dependent on initial efficacy ($l_{s\vartheta}$) and changes in this efficacy over time ($l_{s\gamma}$). Similarly, the proportion of mosquitoes feeding ($k_s$) and being deterred ($m_s$) is dependent on initial impact ($k_{s\vartheta}$, $m_{s\vartheta}$, respectively) and changes over time ($k_{s\gamma}$, $m_{s\gamma}$, respectively).

$$l_s = \frac{1}{1 + \exp\left(-\left(l_{s\vartheta} + l_{s\gamma}\ x\ t\right)\right)'}$$

$$k_s = \frac{1}{1 + \exp\left(-\left(k_{s\vartheta} + k_{s\gamma}\ x\ t\right)\right)'}$$

$$m_s = \frac{1}{1 + \exp\left(-\left(m_{s\vartheta} + m_{s\gamma}\ x\ t\right)\right)'}$$

The percentage of mosquitoes that are repelled without being killed or successfully feeding is represented by $j_s = 1 - l_s - k_s$. The probabilities that a mosquito will feed or will die are conditional on mosquitos not first being deferred before entering a room that has previously received IRS. Here, $k_s$, $l_s$, and $j_s$ are now modified by $m_s$, the degree of deterrence:

$$l'_s = l_s\ x\ (1 - m_s)$$

$$k'_s = k_s\ x\ (1 - m_s)$$

$$j'_s = j_s\ x\ (1 - m_s) + m_s$$

The equations representing the per feeding attempt probability that a mosquito entering a sprayed room is repelled, dies, or survives and feeds, then become:

$$r_s = \left(1 - \frac{k'_s}{k_0}\right) x \left(\frac{j'_s}{l'_s + j'_s}\right)$$

$$d_s = \left(1 - \frac{k'_s}{k_0}\right) x \left(\frac{l'_s}{l'_s + j'_s}\right)$$

$$s_s = \frac{k'_s}{k_0}$$

Additional details can be found in the previously published IRS model changes.[13]

As described in the main text, a non-linear relationship between ITN access and ITN usage was used by converting modelled average population ITN usage into the level of ITN distribution needed to meet this population usage with a stated net distribution frequency. This conversion was performed using an existing model fit usage-distribution curves using data from across sub-Saharan Africa with a Loess curve.[14] ITN access was defined as:

$$ITN\ access = \frac{ITN\ usage}{ITN\ use\ rate}$$

General usage to distribution trends were extrapolated for high access levels without data and the minimum observed values for nets per capita were assigned to all low access levels without data.

ITN access (nets per capita) was converted into nets distributed per person-year, accounting for net retention over time and assuming a distribution frequency of every 3 years (**Table S5**). Population-level loss of ITNs followed a smooth compact function after distribution, where the proportion of nets retained over time, $p(t)$ was represented by:

$$p(t) = \begin{cases} e^{\kappa - \kappa / 1-(\frac{t}{\tau})^2} & if\ t < \tau \\ 0 & if\ t \geq \tau \end{cases}$$

$\kappa$ is a fitted rate parameter estimated from data[14] and $\tau$ represents the time when there are no nets in the population (as estimated from the assumed net half-life):

$$\tau = \frac{ITN\ half - life}{\sqrt{1 - \frac{\kappa}{\kappa - \ln(0.5)}}}$$

The annual nets distributed per capita then equals the integration of the net loss function over a distribution cycle, where $DF$ represents the distribution frequency:

$$Nets\ distributed\ per\ capita\ per\ year = \frac{nets\ per\ capita}{DF * \int_0^{DF} p(t)\ dt}$$

**Table S5**. **Parameter values for the insecticide-treated net (ITN) usage-distribution conversion**.

| Parameter | Symbol | Value | Source |
|---|---|---|---|
| ITN usage | - | Varies in simulations | - |
| ITN use rate (proportion) | - | Varies in simulations | Ranges across African countries by year [14,15] |
| ITN half-life (years) | - | Varies in simulations | Ranges across African countries in 2020 [14,16] |
| ITN distribution frequency (years) | $DF$ | Varies in simulations | World Malaria Report [17] |
| Net loss function rate parameter | $\kappa$ | 20 | Bertozzi-Villa *et al.*, 2021 [14] |

Seasonal malaria chemoprevention

Seasonal malaria chemoprevention (SMC) is implemented in the model as a specified number of rounds of sulfadoxine-pyrimethamine and amodiaquine (SP+AQ), given in consecutive months to children between 3-59 months of age per current World Health Organization recommendations. Doses are timed to overlap with the peak seasonal months of highest malaria transmission intensity.

Similar to routine treatment of clinical infections, SP+AQ directly treats existing infections in the model with a drug-specific probability, moving treated infected individuals to a state of prophylaxis, and providing a period of drug-dependent prophylaxis to uninfected treated individuals.

The probability that an SMC-treated individual is protected from clinical malaria at time $t$, $P_{SPAQ}(t)$, is defined by using a Weibull cumulative distribution function:

$$P_{SPAQ}(t) = \exp^{-(t/\lambda)^k}$$

where $k$ and $\lambda$ are the shape and scale parameters.

SMC parameter values used in the model are included in **Table S3**.

RTS,S/AS01

The RTS,S model was previously fitted to Phase III anti-circumsporozoite antibodies and vaccine efficacy data across 11 sites in sub-Saharan Africa of varying transmission intensity.[18]

The model has two components, antibody dynamics, and efficacy against clinical malaria. The antibody component is comprised of a biphasic exponential decay function. We assume that antibody titers peak $(CSP_{peak})$ following three primary doses of RTS,S, and subsequently decline over time $(t)$ with short-lived $(r_s)$ and long-lived $(r_l)$ decay rates:

$$CSP(t) = CSP_{peak}(\rho_{peak}e^{-r_s t} + (1 - \rho_{peak})e^{-r_l t})$$

$$r_s = log_e(2)/d_s$$

$$r_l = log_e(2)/d_l$$

$d_s$ and $d_l$ represent the half-lives of the short and long-lived cells, respectively. $\rho_{peak}$ and $1 - \rho_{peak}$ represent the proportion of antibodies generated by short-lived and long-lived cells, respectively.

When a fourth dose is given at time $t_{boost}$, titres reach a new peak $(CSP_{boost})$ and subsequently decline. The antibody decay rate remains the same following dose four as that following doses one to three, but the proportion of the response that is generated by short lived cells $(\rho_{boost})$ is modified to simulate different antibody dynamics following a booster dose. Antibody dynamics are described by the following for timepoints following the fourth dose:

$$CSP(t) = CSP_{boot}(\rho_{boost}e^{-r_s(t-t_{boost})} + (1 - \rho_{boost})e^{-r_l(t-t_{boost})})$$

RTS,S parameter values used in the model are included in **Table S3**.

Antibody titers relate to vaccine efficacy against infection over time through a Hill function dose-response curve:

$$V(t) = V_{max} 1 - \left( \frac{1}{1 + \left( \frac{(CSP(t)}{\beta} \right)^{\alpha}} \right)$$

$V_{max}$ represents the maximum efficacy, and $\alpha$ and $\beta$ are shape and scale parameters, respectively.

**Metapopulation model**

The *malariasimulation* metapopulation model extends the transmission model described above to allow simultaneous interaction and modelling between multiple units (or iterations of the standard *malariasimulation* model). The metapopulation model can be used to model a heterogenous region or group of administrative units simultaneously, allowing each to influence the others at every timestep. This process simulates interactions such as human movement across borders. Rather than model individuals in the model moving from one unit to the other, influence of units upon one another is captured through a mixing matrix which allows the EIR and the FOIM of each unit to influence the EIR and FOIM of neighboring units.

<u>Mixing matrices</u>

To capture the connectedness between each of the administrative units in each cluster, we constructed a mixing matrix where each row of the mixing matrix represents the origin administrative unit (termed the primary unit), and each column represents all other administrative units in the cluster, termed secondary units. Each element of the mixing matrix is the normalized time that an individual who normally resides in location $i$ spends in location $j$. To capture this connectivity, we focus only on the movement of people between these locations (i.e. we ignore local movement of mosquitoes).

Transmission between the administrative units is governed by the action that this mixing matrix has on the EIR (i.e. the probability that a susceptible person becomes infected) and on the force of infection acting on mosquitoes (FOIM, the probability that a susceptible mosquito becomes infected). Each row of the matrix sums to 1 to represent all travel originating from each administrative unit. Matrices can be asymmetrical, meaning that the probability of travel from A -> B is not required to equal the probability of travel from B -> A.

To capture movements that occur on a much finer grid than first level administrative units, we constructed some components of the matrix on a 0.1x0.1 degree surface for each cluster set. The population of each cell was summed within the grid boundaries and travel times between each cell were calculated between the cell centroids. After additional calculations described below, this matrix was then aggregated to the first level administrative unit for input into the simulation model.

To construct the matrix, we require three pieces of information:

a. The probability that an individual makes a trip outside their home location in the last 12 months (focusing on overnight stays) and the average number of trips made,
b. The destination of each trip, and
c. The average duration of each trip (in days).

Let $q(i)$ denote the probability that an individual residing in location $i$ makes a trip in the last 12 months, $m(i)$ the average number of trips made and $u(i,j)$ the average duration of a trip to destination $j$. Then the average number of nights that an individual from location $i$ spends outside their home in location $j$ is:

$$a(i,j) = q(i)m(i)u(i,j)$$

Let $P(j|i)$ denote the probability that these trips are made to location $j$. Then the number of nights spent by an individual from location $i$ in location $j$ is:

$$a(i,j)P(j|i)$$

The number of nights spent not travelling (i.e. in their home location $i$) is:

$$365 - \sum_{j \neq i} a(i,j)P(j|i)$$

The non-normalized mixing matrix, $M(i,j)$, is thus given by:

$$\begin{array}{ll} M(i,j) = 365 - \sum_{j \neq i} a(i,j)P(j|i) & i = j \\ a(i,j)(P(j|i) & i \neq j \end{array}$$

The mixing matrix is then normalized by dividing by 365 such that the row sums equal 1 to give the probability that an individual is in each location on any given night.

$u(i,j)$ and $P(j|i)$ were constructed using a 0.1x0.1 degree grid and then aggregated to the first administrative unit level, whereas $q(i)$ and $m(i)$ were constructed at the DHS cluster level and then aggregated to the first administrative unit level.

*a. Probability of travel outside the home location*

We obtained estimates of the probability of making an overnight trip from Demographic and Health Survey (DHS) data on the number of trips away from home for one or more nights in the last 12 months, taken from individual recode and male recode country datasets.[19] Survey results were summarized across sex and year. Data from all available DHS for the countries included in the analysis were pulled and grouped by the first administrative unit level by assigning geolocated clusters to unit shapefiles. The median number of overnight trips was calculated at the DHS cluster level and then averaged at the first administrative unit level across sex and survey years. Units with missing data were assigned the mean country level value and countries with missing data were assigned the mean value across sub-Saharan Africa (**Figure S8**). The probability of overnight travel during the last 12 months at the unit level ranged from 0.08 to 0.75 with the mean value across sub-Saharan Africa equal to 0.43. The number of overnight trips in the last 12 months at the unit level ranged from 1.14 to 18 with the mean value across sub-Saharan Africa equal to 2.79.

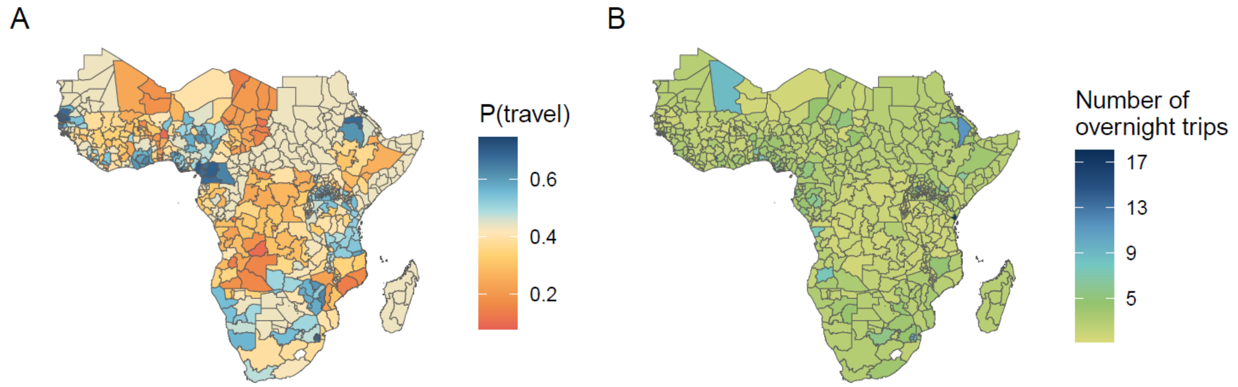


**Figure S8**. Data from the Demographic and Health Surveys Program showing the A) probability of any overnight travel in the last 12 months and B) the number of overnight trips in the last 12 months (admin1 mean of the cluster medians).

*b. Trip destinations*

Let $P(j|i)$ denote the probability that this trip is made to location j. To obtain these values we adapted a previously published gravity model[20] fit using traveler survey data from Burkina Faso, Mali, Tanzania, and Zambia.[21] The gravity model calculates the probability of travel as:

$$P(j|i) \propto N_j^{\tau} k(d_{i,j})$$

where $i$ represents the origin location, $j$ represents the destination location, $N_j$ is the population size at the destination, $\tau$ is a power parameter, and $k(d_{i,j})$ is a distance kernel calculated as:

$$k(d_{i,j}) = \left(1 + \frac{d_{i,j}}{\rho}\right)^{-\alpha}$$

where $\rho$ is a scale parameter and $\alpha$ is a power parameter.

Distance in the original model fits was measured as the Euclidean distance in km from origin to destination point, assuming that travelers move in a straight line at a constant pace per km from origin to destination. We relax this assumption by using an algorithm to calculate the path-of-least-resistance from origin to destination across a friction surface[2] taken from the Malaria Atlas Project. The friction surface uses data from 2015, where each pixel covers 1km x 1km and represents the time needed to travel across the pixel as categorized by water, road, railroad, and land cover datasets.

The gravity model parameters were re-fit for the current analysis using travel time from origin to destination instead of Euclidean distance. Model parameters were fit to the original traveler survey data from Burkina Faso, Mali, Tanzania, and Zambia.[20] Parameters for both fits are given in **Table S6**. Both models were fit using MATLAB 9.14.0 (R2023a) (The MathWorks Inc., 2023). We calculated gravity model estimates on the 0.1x0.1 degree grid by summing the population of each cell within the grid boundaries and calculating travel times between cell centroids. Gravity model values were then aggregated up to the first level administrative unit by summing estimates of all interactions between units.

**Table S6**. **Gravity model parameters (median values with 95% credible intervals)**. Parameters were fitted to origin-destination pairs using survey data collected from Burkina Faso, Mali, Tanzania, and Zambia.

| $\alpha$ | $log(\rho)$ | $\tau$ | **Source** |
|---|---|---|---|
| 1.91 (1.78-2.06) | 4.29 (4.09-4.48) | 1.22 (1.20-1.24) | Marshall et al. 2018 [20] |
| 2.67 (2.47-2.93) | 5.11 (4.94-5.30) | 1.13 (1.11-1.15) | Model fit for this paper |

*c. Trip durations*

Trip durations were calculated using the traveler survey data from Burkina Faso, Mali, Tanzania, and Zambia mentioned above. [21] Quantile regression models were fit to the survey data to estimate the association between trip duration and travel time. The best fitting model as measured by the AIC was a quadratic model (**Figure S9**). Using the model results, we predicted estimated trip durations of trips between each centroid on the 0.1x0.1 degree grid and aggregated up to the first level administrative level, by finding the mean duration of travel between unit interactions.

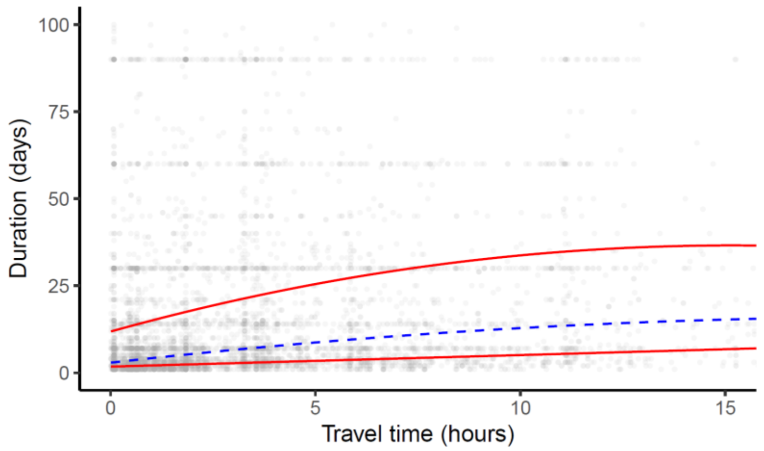


**Figure S9**. **Model fits between trip duration (days) and travel time (hours)**. Each trip is represented by a single gray dot; origin-destination pairs are taken from survey data collected from Burkina Faso, Mali, Tanzania, and Zambia.[21] Models shown are quadratic quantile regression models; the blue dashed line shows the median, and the two red lines show the 25th and 75th percentiles. The figure is cropped to display the bulk of the data, however, maximum trip duration exceeded one year, and maximum travel time exceeded 30 hours.

A three-unit metapopulation example is shown in **Figure S4**, with three levels of mixing and resulting *Pf*PR$_{2\text{-}10}$ estimates across time.

Model dynamics

The mixing matrices described above indicate the probability of travel at any given time step from one unit to all other units in the model run. The mixing matrices are assumed to remain constant in simulations. Rather than modeling individual movement between model units, high- and low-transmission units influence each other through the EIR and the FOIM. The EIR is influenced by travel from origin $i$ to destination $j$, and the FOIM is influenced by travel from destination $j$ to origin $i$. Here $\varepsilon$ represents EIR, $\Lambda_M$ is the force of infection on mosquitoes, $i$ is the unit of interest, $j$ represents all other units in the metapopulation model $t$ indicates time, and $\varsigma$ indicates the probability of travel between units as drawn from the specified mixing matrix:

$$\varepsilon_i(t+1) = \varsigma_{i,j}\varepsilon_i(t) + \sum_j \varsigma_{i,j}\varepsilon_j(t)$$

$$\Lambda_{Mi}(t+1) = \varsigma_{j,i}\Lambda_{Mi}(t) + \sum_j \varsigma_{j,i}\Lambda_{Mi}(t)$$

$$\sum \varsigma_i = 1$$

Interventions

Border post test-and-treat interventions are modelled by adjusting the effects of mixing between units by a value dependent on the probability of capturing hypothetical border crossings, the probability of identifying infections accurately through RDT, and the probability that treatment successfully clears infection.

$$coefficient = 1 - P(checked) \times P(RDT_+ \cap\ infected) \times P(treated)$$

Here, $P(checked)$ is the proportion of border crossings that are captured by the intervention. A value of 1 means that 100% of travelers are checked at the border post, a value of 0 means that no travelers are checked.

$P(RDT_+ \cap\ infected)$ is the probability that travelers from the source unit are infected and would be detected as a case by a rapid diagnostic test (RDT). We assume that RDT specificity is high and therefore approximate the probability of true positives with RDT prevalence, $P(RDT_+ \cap\ infected) \approx P(RDT_+)$. We derive an RDT prevalence from PCR prevalence, assumed to be the proportion of individuals in the $D$, $A$, and $U$ (diseased, asymptomatic, and sub-patent) infection states, and convert it to RDT prevalence using fits taken from previously published parameters[3,4] (**Figure S5**).

$P(treated)$ is the probability treatment would be successful. In this analysis, treatment is with AL which is assumed to successfully clear infection in 95% of individuals.[5]

The calculated $coefficient$ for each setting is then added into the equation to calculate mixed EIRs:

$$\varepsilon_i(t+1) = \varsigma_{i,i}\varepsilon_i(t) + \sum_j \varsigma_{i,j}\varepsilon_j(t) \ x \ coefficient$$

$$\sum \varsigma = 1$$
